# Identification and Quantification of Autoantibodies against Prostate-Specific Antigens by Immunoaffinity-Mass Spectrometry

**DOI:** 10.1101/2025.06.26.25330371

**Authors:** Yasmine Rais, Andrei P. Drabovich

## Abstract

Prostate-specific antigen (PSA) utilized clinically to diagnose prostate cancer (PCa) has limitations of low diagnostic specificity and lack of prognostic information. Novel PCa markers are needed to improve PCa diagnostics. Here, we hypothesized that some prostate-specific antigens leaking into systemic circulation during prostate tissue transformation and PCa progression could trigger production of autoantibodies, and that these autoantibodies could improve PCa diagnostics. Autoantibodies against prostate-specific antigens were previously exclusively detected by serological immunoassays, but the existence of autoantibodies was often debated due to potential cross-reactivity and non-specific binding of indirect immunoassays. Here, we aimed at developing a proteome-wide platform for serological assays that could discover and quantify antigen-specific autoantibodies in blood serum and evaluate their diagnostic potential. We developed targeted and shotgun Immunoaffinity-Mass Spectrometry (IA-MS) assays to quantify serum autoantibodies against prostate-specific proteins PSA (kallikrein-3; KLK3_HUMAN), kallikrein-4 (KLK4_HUMAN), prostate-specific membrane antigen (FOLH1_HUMAN), and prostatic acid phosphatase (PPAP_HUMAN). IA-SRM assays resolved false positive identifications, discovered IgG1, IgA1, and IgM as the most prevalent isotypes of autoantibodies, and provided reproducible quantification of autoantibodies in negative biopsy, low-risk PCa, and metastatic PCa serum samples. Anti-KLK3 and anti-KLK4 IgG1 autoantibodies were detected in 75% (median concentration 4 ng/mL) and 67% (median 11 ng/mL) of PCa serum samples, respectively. Indirect immunoassays collectively detecting IgG autoantibodies, a mixture of four subclasses, revealed poor signal-to-noise ratios, false positives, and a surprisingly high number of false negatives, debating the usefulness of indirect immunoassays for discovery and quantification of autoantibodies. The presented proteome-wide serology assays will facilitate the quantification of PCa autoantibodies, paving the way to improved diagnostics of PCa and comprehensive evaluation of immune response to prostate-specific antigens.

## INTRODUCTION

Prostate cancer (PCa) is the third leading cause of cancer mortality in men^1^. Prostate-specific antigen (PSA), a widely used marker for PCa management, has low diagnostic specificity and lacks prognostic value; this often results in over-diagnosis and over-treatment of low-risk PCa^2–7^. Some PCa patients, however, are initially diagnosed with or later progress to the high-risk PCa and then to metastatic castration-resistant prostate cancer (mCRPC)^8^. With the limited treatment options of mCRPC and short survival advantage, mCRPC remains an incurable disease^9^.

Immunotherapy has recently revolutionized the treatment of metastatic cancers^10^, but not mCRPC^11–14^. The most advanced autologous cellular vaccines targeting prostate-specific proteins KLK3_HUMAN (PROSTVAC-VF) and PPAP_HUMAN (Sipuleucel-T) showed only marginal survival advantage^8^. Additional prostate-specific proteins (FOLH1_HUMAN^15^, TGM4_HUMAN^16^, KLK4_HUMAN^17^) were found immunogenic, but need to be evaluated for their potential as novel immunotherapy targets.

With the mentioned limitations of PSA, there has been intense work on the identification of novel PCa biomarkers^18^ and immunotherapy targets^9,19^. Genetic markers were studied to evaluate PCa predisposition^20–24^. Tissue-based and cell-free DNA gene expression panels such as Decipher, Oncotype Dx, ConfirmMDx, and others were introduced for PCa risk stratification on biopsy^25^. RNA markers in urine were among the most developed and included PCA3-based Progensa^26,27^, TMPRSS2-ERG^28^, and several panels^25^. Protein markers were extensively investigated^29–31^ including our studies on KLK4^32^, TGM4^33^, RLN1^34^, and TMPRSS2-ERG protein^35^. Several PSA forms, such as total, free, intact, and [-2]proPSA, alone (Prostate Health Index) or in combination with KLK2 (4K score), emerged as FDA-approved blood-based panels for consideration of initial or repeated biopsies in patients with elevated PSA^25^. None of these other markers has yet been proven to predict high-risk PCa based on minimally invasive blood or urine tests.

Over the past decades, autoantibodies have been identified in the serum of patients with many cancers^36,37^. Several autoantibody panels (Videssa Breast^38^, EarlyCDT-Lung^39^, Mitogen Dx cancer^40^) are now commercially available. Autoantibodies were often detected at early stages before presentation of clinical symptoms, providing a basis for early cancer diagnosis^41^. Previous studies have shown that a combination of PSA levels with anti-PSA autoantibody levels could increase diagnostic specificity to distinguish PCa from noncancerous prostate enlargement such as benign prostatic hyperplasia (BPH)^42^. Conversely, the presence of anti-PSA autoantibodies could interfere with PSA testing and yield significantly falsified results of PSA testing^43^.

Previous autoantibody studies, however, had limitations: autoantibodies were exclusively detected and measured by indirect immunoassays and have never been validated by alternative analytical techniques. Indirect immunoassays were prone to false-positive detection^34,44^, provided semi-quantitative relative measurements, and lacked standardization and multiplexing for isotypes and subclasses. As a result, the phenomenon of autoantibodies often raised skepticism in cancer research.

Recently, we developed a proteome-wide platform for serological assays that could unambiguously discover and quantify antigen-specific endogenous human antibodies^44–47^. Immunoaffinity-shotgun Mass Spectrometry (IA-MS) assays revealed proteome-wide diversity of isotypes (IgG, IgM, IgA, IgD, IgE), subclasses (IgG1, IgG2, IgG3, IgG4, IgA1, and IgA2) and Fc-interacting proteins (C1q complex components) which were enriched by viral antigens, with the distinct infectious-specific differences^44,46^. Simple design and execution of our targeted immunoaffinity-selected reaction monitoring (IA-SRM) assays empowered with carefully-designed synthetic peptide internal standards provided robust, selective, sensitive (1 ng/mL), reproducible, and high throughput (140 samples/day) assays devoid of limitations of indirect immunoassays^44,46^.

In this study, we developed and applied IA-MS assays (**Figure 1**) to discover and quantify serum autoantibodies against prostate-specific proteins PSA (kallikrein-3; KLK3_HUMAN), kallikrein-4 (KLK4_HUMAN), prostate-specific membrane antigen (FOLH1_HUMAN), and prostatic acid phosphatase (PPAP_HUMAN), and evaluate diagnostic potential of these autoantibodies.

**Figure 1.**
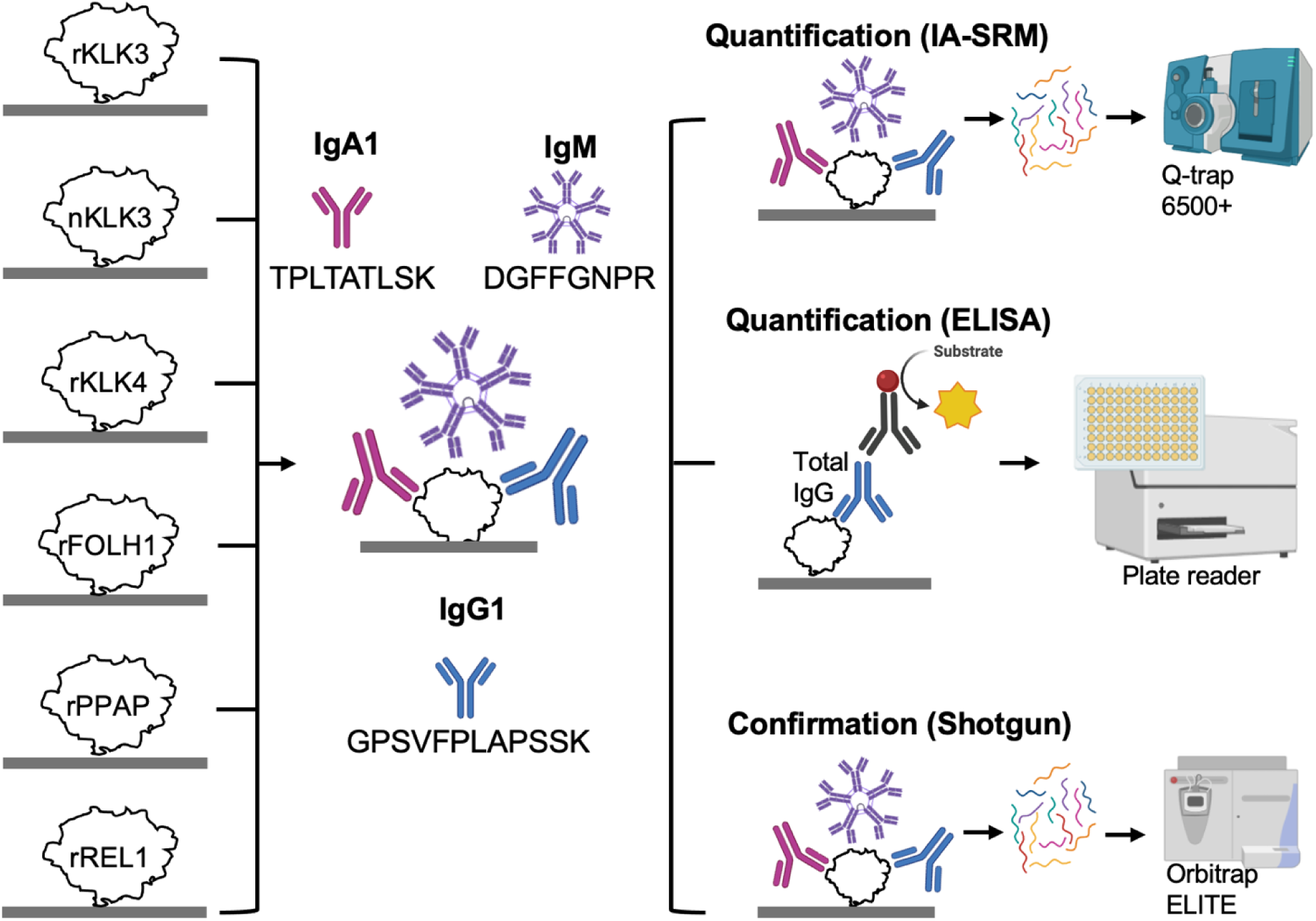
Experimental workflow for the discovery and quantification of PCa autoantibodies. Prostate tissue-specific proteins PSA (kallikrein-3; KLK3_HUMAN), kallikrein-4 (KLK4_HUMAN), prostate-specific membrane antigen (FOLH1_HUMAN), and prostatic acid phosphatase (PPAP_HUMAN) were evaluated as antigens. REL1_HUMAN was a negative control, and two forms of PSA were evaluated: recombinant (rKLK3) and native endogenous PSA purified from human seminal fluid (nKLK3). IA-SRM assays were used for the quantification of the complete panel of human immunoglobulin isotypes and subclasses (IgG1, IgG2, IgG3, IgG4, IgA1, IgA2, IgM, IgD, and IgE), while indirect ELISA measured antigen-specific IgG, a mixture of four subclasses. IA-shotgun MS/MS was utilized for independent confirmation of immunoglobulin subclass identity and their further characterization, such as discovery of secreted or membrane isoforms.

## EXPERIMENTAL PROCEDURES

### Hypothesis, Study Design, and Objectives

We hypothesized that some prostate tissue-specific antigens leaking into systemic circulation during prostate transformation and PCa progression could trigger production of autoantibodies, and that these autoantibodies could improve PCa diagnostics. We designed a retrospective study with the clinically defined cohorts of biobanked blood serum samples. Two orthogonal assays were used: (i) a conventional indirect ELISA immunoassay for relative quantification, and (ii) a novel IA-SRM assay for absolute quantification of autoantibody levels in serum. Our study included the following objectives: (i) confirm the existence of autoantibodies against prostate-specific proteins; (ii) demonstrate the advantages of IA-SRM assay versus indirect immunoassays; (iii) distinguish between negative biopsy and PCa based on autoantibody profiles; (iv) evaluate correlations between levels of PSA and anti-PSA autoantibodies; (v) prioritize antigens with diagnostic potential; (iv) overview implications of detected autoantibodies for molecular diagnostics, diagnostic imaging, and PCa immunotherapy.

### Chemicals and reagents

Dithiothreitol, iodoacetamide, and trifluoracetic acid (TFA) were purchased from Thermo Fisher Scientific (Burlington, ON). Mass spectrometry-grade Optima acetonitrile (ACN) and water were obtained from Fisher Scientific (Fair Lawn, NJ). Formic acid (FA), and dimethylated SOLu-trypsin were obtained from Sigma-Aldrich (Oakville, ON).

### Proteins, antibodies, and peptide standards

An indirect immunoassay detection antibody (goat-anti-human IgG Fcγ conjugated to horseradish peroxidase; A18817) was obtained from Invitrogen. Recombinant human antigens KLK3_HUMAN (10771-H08H), KLK4_HUMAN (11857-H08H), FOLH1_HUMAN (15877-H07H), PPAP_HUMAN (10959-H08H), and REL1_HUMAN (11625-H08H) were obtained from Sino Biological (**Supplementary Table S1**). Native PSA protein from human seminal fluid (#497-11-0.1) was obtained from Medix Biochemica (Finland). Synthetic stable isotope-labeled peptide internal standards with trypsin-cleavable and quantifiable tags (SpikeTides_TQL) were provided by JPT Peptide Technologies (Germany).

### Clinical samples

Thirty blood serum samples were obtained from patients with biopsy-confirmed PCa (15 low-risk PCa and 15 mCRPC), and 20 blood serum samples were obtained from patients with negative biopsy, a noncancerous enlargement of the prostate such as benign prostatic hyperplasia (**Supplementary Table S2**). Blood serum samples were obtained from the Alberta Prostate Cancer Registry & Biorepository. The study was approved by the Health Research Ethics Board of Alberta (HREBA) Cancer Committee (HREBA.CC-22-0056 and HREBA.CC-14-0085).

### ELISA

The recombinant proteins rKLK3 (PSA), rKLK4, rFOLH1, rPPAP, and rREL1 were diluted in PBS (pH 7.4), individually coated (0.2 μg/well; 100 μl/well) onto high-binding 96-well polystyrene microplates (07000128; Greiner Bio-One) and incubated overnight at room temperature (RT). After washing with 0.1% Tween-20 in PBS (washing buffer; three times with 200 μl), plates were blocked with a blocking buffer (6% BSA in PBS) for 1-hour at RT. Blood serum samples were diluted in blocking buffer (1:80) and added to plates (100 μL per plate). After 2 h incubation with continuous shaking, plates were washed three times with 200 μl washing buffer, and the detection antibody (HRP-conjugated goat-anti-human IgG Fcγ; 0.02 μg/μL) was added and incubated for 1 h at RT. After final washing, 100 μL of tetramethylbenzidine (TMB) solution was added, the reaction was stopped after 5 min with 50 μL of 2 M HCl. Absorbance (OD 450 nm) was measured using FilterMax F5 multi-mode microplate reader (Molecular Devices) with 450NMBW80 absorbance filter.

### Immunoprecipitation and proteomic sample preparation

To measure PCa autoantibodies by IA-MS, high-binding 96-well polystyrene microplates (#07000128; Greiner Bio-One) were coated overnight with recombinant proteins rKLK3, rKLK4, rFOLH1, rPPAP, or rREL1 (0.3 μg/well), blocked for 1 h with blocking buffer (5% sucrose, 50 mM Tris-HCL, 500 mM potassium chloride, 0.05% Tween-20 diluted in PBS; pH 7.6), and washed. Blood serum samples (20 μl) were diluted 1:5 with the blocking buffer, and 100 μL were added per well. Samples were incubated with continuous shaking for 2 h at RT, and microplates were washed three times with a washing buffer (200 μl) and three times with 50 mM ammonium bicarbonate (200 μl). Enriched autoantibodies and proteins were subjected to proteomic sample preparation directly on the same plate used for IA enrichments. SpikeTides_TQL (**Supplementary Table S3**) peptide standards (500 fmol/well) were added before proteomic sample preparation. Autoantibodies and proteins were denaturated, and disulfide bonds were reduced by 10 mM dithiothreitol at 70 °C for 15 min and alkylated with 20 mM iodoacetamide at room temperature (RT) in the dark for 45 min. The denatured and alkylated autoantibodies and proteins were diluted to 110 μl with Milli-Q water. On-plate digestion was completed overnight at 37 °C using recombinant dimethylated SOLu-trypsin (0.2 μg/well). Finally, the digestion was stopped with trifluoroacetic acid, transferred with a multichannel pipette onto 96-well plates compatible with HPLC autosamplers (full skirt Axygen PCR plates; AXYPCR96FSCS; Sigma-Aldrich), sealed with silicone mats, and stored at -20° C until use.

### Rapid IA-HPLC-SRM assays

Plates with sample digests were thawed and centrifuged to remove air bubbles; 30 μL of digest was injected per technical replicate. Peptides were desalted, washed and concentrated onto a Luna C18 trap column (30 mm × 1 mm; 3 μm 100 Å; #00A-4251-A0; Phenomenex, Torrance, CA) at a flowrate of 170 μL/min, and separated at a flowrate of 40 μL/min on a Luna Omega C18 analytical column (50 mm × 1 mm, 1.6 μm, 100 Å, 25°C; #00B-4748-A0). A 6-port diverter valve supplied with QTRAP 6500+ was used to divert the flow either into waste during loading onto trap column, or onto analytical column during peptide separation and data acquisition. The gradient started with 95% buffer A (0.1% formic acid in water) and 5% buffer B (0.1% formic acid in acetonitrile) for 2 min and ramped to 25% over 0.7 min, 37% over 4.1 min, 44% over 0.6 min, 95% over 1.2 min, 5% over 0.3 min, and continued for 1.1 min. High-performance UPLC (Waters Acquity Classic), optiFlow Turbo V nanospray ion source, 50 μl ESI probe, and QTRAP 6500+ (Sciex, Concord, ON) ensured highly sensitive and fast (10 min analysis time per injection) quantification of autoantibodies. QTRAP 6500+ acquisition parameters were: 4700 V ion spray; 300° C source temperature; 35 psi curtain gas; 18 psi for atomizing Gas 1 and 15 psi auxiliary Gas 2; 80 V declustering potential; 10 V entrance potential, 12 V collision cell exit potential, and collision gas “High”. The detailed SRM parameters are presented in **Table 1**. Each patient sample was measured with one analytical (“full process”) replicate and two technical replicates. The third technical replicate was preserved as a backup.

**Table 1.**
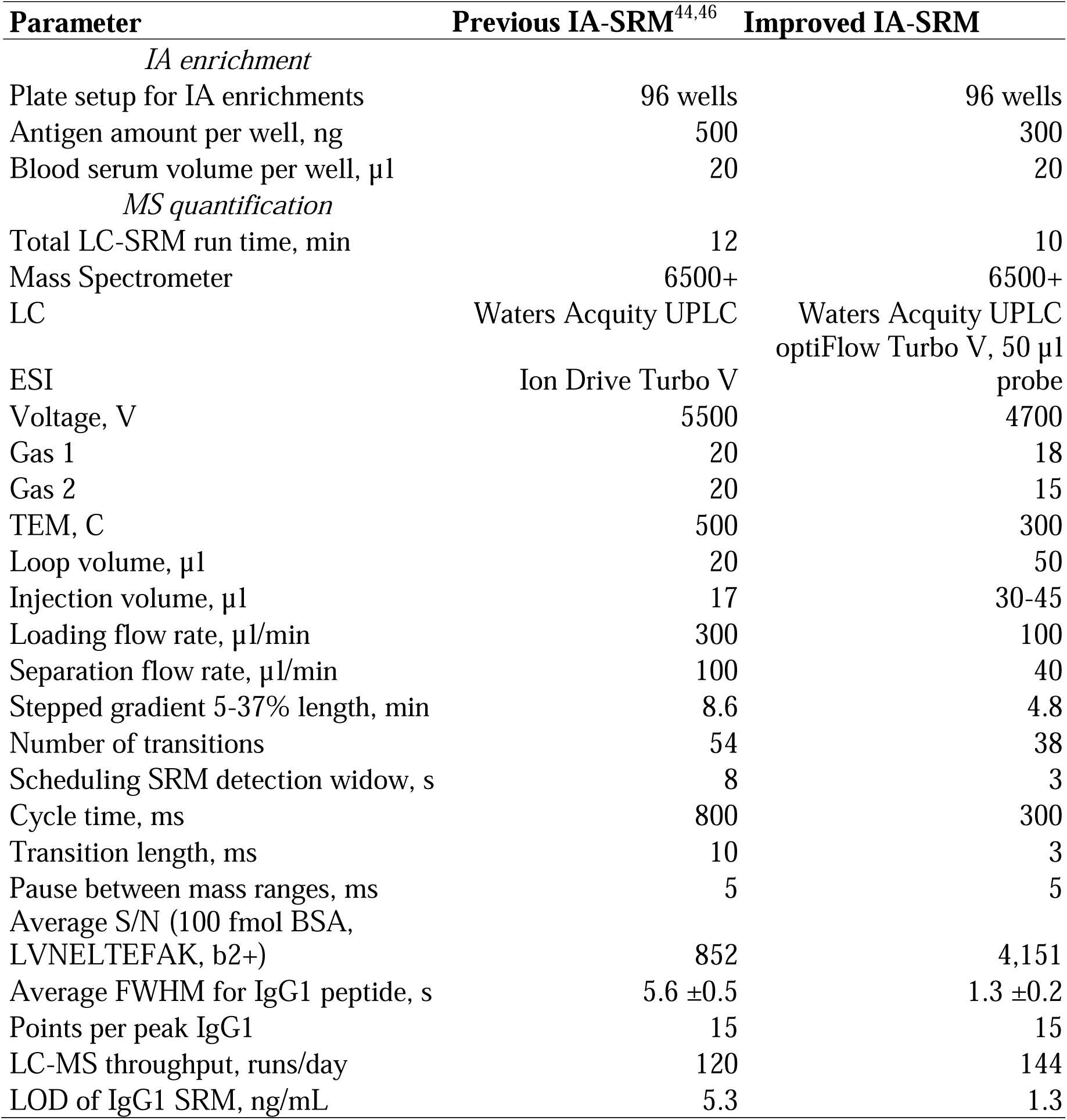
Parameters of an improved IA-SRM assay for quantification of endogenous autoantibodies.

### Shotgun LC-MS/MS

Autoantibody isotypes and subclasses were identified with nanoflow liquid chromatography-tandem mass spectrometry (nLC–MS/MS) using Orbitrap Elite mass spectrometer equipped with an Easy-Spray source, Nano EASY-Spray emitter (ES993), and an EASY-nLC II system (Thermo Scientific). OMIX C18 10 μl tips (Agilent Technologies) were used for desalting and microextraction of tryptic peptides. A trap column (μPAC trapping column; COL-TRPNANO16G1B2, Thermo Scientific) was used for sample loading at a flowrate of 15 μl/min, while analytical column (110 cm μPAC Neo; COL-NANO110NEOB, Thermo Scientific) was used for peptide separation at 400 nL/min. Peptides were separated with the following gradient: 95% buffer A (0.1% formic acid in water) and 5% buffer B (0.1% formic acid in ACN) at 0 min, 5–20% B for 30 min, 20–30% B for 5 min, 30–45% B for 2 min, 30–90% B for 2 min, and 90% B for 10 min. Orbitrap FTMS full scans (303–1200 m/z) were performed at 60 K resolution (at 400 m/z; lock mass ion 445.120025) in the profile mode, followed by 11 Data Dependent Acquisition FTMS MS/MS scans in Orbitrap (15 K at 400 m/z; centroid mode; 2 m/z isolation width, 0.1 ms activation time; 30% normalized collision energy; 200-1600 m/z range; monoisotopic precursor selection; unassigned and +1 charge states rejected; dynamic exclusion for 11 s). Minimal signal threshold for MS/MS was lowered to 3,000 counts. FTMS full AGC and MSn AGC targets were set to 1 × 10^6^ (140 ms injection time) and 40,000 (140 ms injection time), respectively. Instrument parameters included 290 °C capillary temperature, 2.4 kV source voltage, and 69% S-lens RF level.

### MS Data Analysis

QTRAP 6500+ raw files were acquired with Analyst software (v1.7.3; Sciex) and analyzed using Skyline Targeted Proteomics Environment v20.1.0.76^48^. The peak boundaries were adjusted manually, and the integrated areas of all transitions for each peptide were extracted. Light-to-heavy peak area ratios were used for absolute quantification of endogenous peptides. IgG and IgA antibodies were considered as monomers (2 copies of internal standard peptides per IgG or IgA), while IgM was considered as a pentamer (10 peptide copies per IgM). Orbitrap Elite raw files were acquired with XCalibur software (v2.2) and searched with MaxQuant v2.6.1^49^ against the reviewed human canonical Uniprot database (2022-07-03; 20,286 proteins) which was modified to include several isoforms of human immunoglobulin heavy constant genes, secreted isoforms, as well as secreted isoforms missing lysine residues at the immunoglobulin heavy chain C terminus. Search parameters included: trypsin enzyme specificity, two missed cleavages, 6 aa minimum peptide length, 20 ppm first search and 4.5 ppm main search MS1 match tolerance, and 20 ppm FTMS MS/MS match tolerance. Modifications included fixed cysteine carbamidomethylation and variable methionine oxidation, protein N-terminal acetylation, and asparagine deamidation. False-discovery rates for peptide and protein identifications were set to 1.0%. Label-free quantification (LFQ) algorithm was used for protein quantification.

### Experimental Design and Statistical Rationale

In an indirect ELISA study, serum samples were enriched with a specific antigen (N=50 samples per antigen; 4 antigens) and a no-antigen control (PBS; N=50 samples). Thus, 250 samples were analyzed with a single analytical and a single technical replicate. In an IA-SRM study, serum samples were analyzed with the single-point titration with 500 fmol/well of SpikeTides_TQL heavy peptide internal standards, single analytical replicate, and two technical replicates per sample. Our rationale to measure single analytical replicates was explained by the large inter-patient variability of autoantibody levels (CV∼50%) substantially exceeding analytical and technical reproducibility of ELISA measurements and IA-SRM measurements with SpikeTides_TQL internal standards (CV<10%). In the discovery phase, we aimed at providing evidence that the levels of IgG or IgG1 autoantibodies enriched with specific antigens would be significantly higher than non-specific IgG or IgG1 levels enriched with negative control antigens (PBS or REL1 protein) from the matched serum samples and parallel enrichment and sample preparation. As an evidence cut-off, we have chosen a widely accepted cut-off for autoantibody immunopositivity calculated as a mean plus two standard deviations^50,51^. Indirect ELISA measurements of IgG non-specifically enriched from serum in PBS-coated wells (negative controls) revealed 0.61 au mean optical density, 0.34 au standard deviation, and 56% CV (N=50 serum samples). An immunopositivity cut-off (1.29 mean, 56% CV, 0.72 standard deviation) would result in 1.09 effect size and would require at least 9 matched serum samples (positive or negative antigen enrichments) to achieve power >0.8 (two-tailed *t*-test for matched pairs; α = 0.05; G∗Power software, v3.1.7, Heinrich Heine University Dusseldorf). IA-SRM measurements of IgG1 non-specifically enriched from serum in PBS-coated wells (negative controls) revealed 1.7 ng/mL mean concentration, 0.5 ng/mL standard deviation, and 32% CV. An immunopositivity cut-off (2.8 ng/mL mean concentration, 32% CV, 0.9 ng/mL standard deviation) would result in 1.4 effect size and would require at least 7 matched serum samples (positive or negative antigen enrichments) to achieve power >0.8 (two-tailed *t*-test for matched pairs; α = 0.05). To evaluate diagnostic utility of autoantibodies for distinguishing between negative biopsy and PCa serum, we assumed that IgG or IgG1 autoantibodies would be elevated ≥2-fold in PCa. To detect with 0.8 power autoantibodies elevated ≥2-fold by ELISA (effect size 1.13; inter-patient CV=56%; α=0.05; allocation ratio 1.5; 2-tailed MWU), we need to measure autoantibodies in at least 12 negative biopsy and 18 PCa samples. To detect with 0.8 power autoantibodies elevated ≥2-fold by IA-SRM (effect size 1.98; inter-patient CV=32%; α=0.05; allocation ratio 1.5; 2-tailed MWU), we need to measure autoantibodies in at least 5 negative biopsy and 7 PCa samples. To summarize, the selected set of samples (30 negative biopsy, 15 low-grade PCa, and 15 mCRPC) was suitable to design a powered study to discover autoantibodies and evaluate their diagnostic utility by either ELISA or IA-SRM assays.

## RESULTS

### Measurement of anti-PSA autoantibodies by indirect immunoassays

Since numerous studies have previously reported the presence of anti-PSA autoantibodies in blood serum of patients with prostate cancer^52–54^, we designed preliminary experiments to detect autoantibodies against a recombinant PSA (rKLK3) using indirect immunoassays. A conventional indirect immunoassay measured anti-rKLK3 IgG without differentiation into individual subclasses of IgG1, IgG2, IgG3, and IgG4. Measurement of anti-rKLK3 IgG autoantibodies in 50 serum samples, including 20 negative biopsy (nBx), 15 low risk cancer (LR-PCa), and 15 mCRPC samples (**Figure 2A**) revealed a relatively high background in no-antigen negative controls (microplates coated with PBS buffer instead of 200 ng of rKLK3 antigen). Similar to previous studies^52–54^, we utilized no-antigen negative controls to derive a cut-off for positive immunoreactivity which was defined as a mean plus two standard deviations of ELISA optical density values. This cutoff revealed a relatively low immunopositivity in 5% of nBx and 17% of PCa samples. Detected immunopositivity was in line with previous data (11% in PCa^54^, 13% in mCRPC^52^, 7% in PCa^52^, 3.5% in PCa^53^).

**Figure 2.**
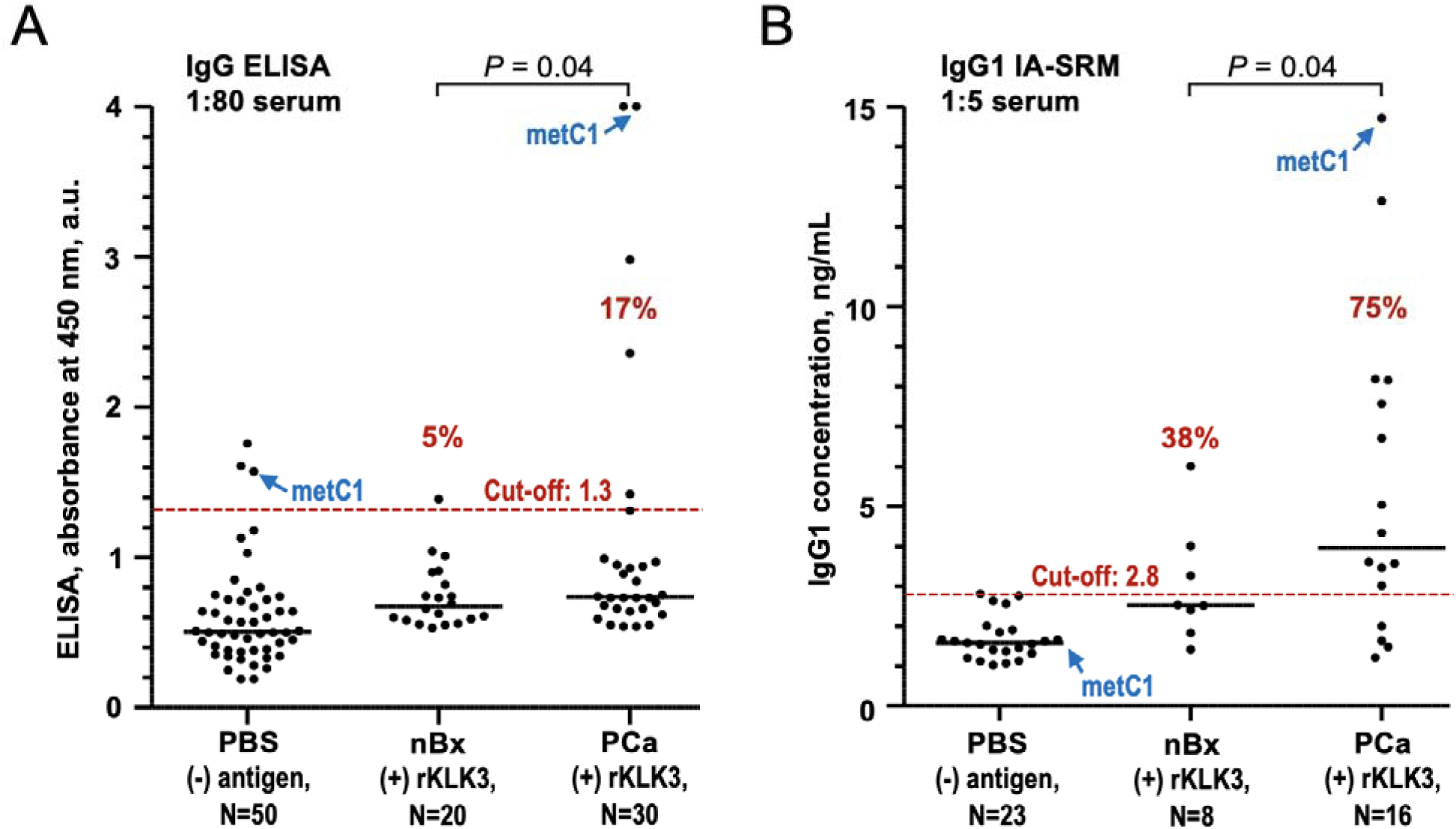
Evidence for serum anti-rKLK3 autoantibodies. (**A**) Indirect ELISA revealed anti-rKLK3 IgG in serum (1:80 dilution) of PCa and nBx patients, with the immunopositivity cut-off of 1.3 calculated as a mean plus two standard deviations in a PBS negative control. Immunopositivity in PCa samples was 17%. Diagnostic performance (PCa vs. nBx) was statistically significant (2-tailed Welch’s t-test *P = 0.040*). (**B**) IA-SRM assay detected anti-rKLK3 IgG1 in serum of PCa and nBx patients (1:5 dilution). Immunopositivity in PCa samples was 75%. Diagnostic performance (PCa vs. nBx) was statistically significant (2-tailed Welch’s t-test *P = 0.040*). A metastatic sample metC1 measured by IA-SRM with and without an antigen revealed signal-to-noise ratio of 8.8, as compared to 2.4 ratio for metC1 measured by ELISA.

### Development of IA-SRM assays for identification and quantification of anti-PSA autoantibodies

We previously demonstrated the utility and advantages of IA-MS and IA-SRM assays to identify and quantify serum antibodies against numerous infectious disease antigens such as SARS-CoV-2 Spike, S1, RBD, and N proteins^44^, and RSV glycoproteins F and G^46^. Stable isotope labeled internal standards used as assay calibrators revealed for the first time the absolute concentrations of anti-viral antibodies, as opposed to the relative values of titers or optical density. Interestingly, convalescent serum revealed relatively high levels of anti-viral IgG1 in the range of 1,000 – 3,000 ng/mL^44,46^, resulting in the high SRM signal intensity and confident reproducible measurements of anti-viral IgG1. Our previous IA-SRM assays had a throughput of 120 runs/day, required no additional offline solid phase extraction (protein digests were directly loaded onto the trap column), and had LOD ∼ 5 ng/mL^44,46^. While some previous studies reported anti-PSA IgG levels at 1,400 ng/mL^55^, our preliminary measurements by IA-SRM demonstrated ∼200-fold lower levels. Such low levels of ∼10 ng/mL were approaching the limits of detection and required re-development of our IA-SRM assay.

Here, we re-developed SRM assay (**Table 1**) to push the limits of LC separation (peak widths ∼1 s) while keeping high throughput of 140 runs a day, LOD of IgG1 LOD ∼1 ng/mL, 15 points per peak, simplicity, reproducibility, and overall robustness. These advancements were achieved with switching to the reverse-phase LC columns of 1 mm internal diameter and fine particles of 1.6 µm, installation of a 50 µl injection loop to enable nearly complete utilization of ∼110 µl digest in 2-3 injections, and other critical adjustments (**Table 1**).

Using shotgun MS and proteome-wide searches, we evaluated the purity of recombinant proteins used as antigens in our assays. In terms of presence of other proteins (HEK293 expression system contaminants, etc.), the high purity was found for recombinant antigens rKLK3 (99% pure), rKLK4 (98%), rFOLH1 (96%), and rPPAP (99.9%). While an endogenous native KLK3 purified from seminal fluid (nKLK3) revealed lower purity (56%), it served as an important control having a native glycosylation pattern of prostate tissues^56^.

### Measurement of anti-PSA autoantibodies by IA-SRM

We selected a set of samples with the highest and lowest fold changes for ELISA signal in (+) rKLK3 versus no-antigen PBS controls and measured autoantibody levels by IA-SRM (**Figure 2B**). Due to the high selectivity of MS, low background, and high sensitivity of IA-SRM, we detected anti-rKLK3 IgG1 in 75% of PCa and 38% of nBx sera (**Figure 2B**). Diagnostic performance (**Figure 2B**; PCa vs. nBx) wa statistically significant (2-tailed Welch’s t-test *P* = 0.040).

### Validation of anti-PSA autoantibodies enriched by recombinant versus native KLK3

Our preliminary IA-SRM assays were developed with a recombinant PSA (rKLK3) expressed in a baculovirus insect system. Since different protein expression systems may lead to different glycosylation profiles and additional non-specific binding, we validated autoantibodies enriched with a native PSA purified from human seminal fluid (nKLK3). As a result, we observed a similar response for anti-PSA IgG1 enriched with rKLK3 and nKLK3 (**Supplementary Figure S1**).

### Discovery of autoantibodies against prostate-specific antigens rKLK4, rFOLH1, and rPPAP

In addition to rKLK3, we measured autoantibodies against other prostate-specific antigens including, KLK4_HUMAN, PPAP_HUMAN, REL1_HUMAN, and FOLH1_HUMAN. Similar to KLK3, KLK4 (kalikrein-4), PPAP (Prostatic acid phosphatase), FOLH1 (prostate-specific membrane antigen) and REL1 (relaxin 1) are the secreted proteins whose transcripts were ranked within top 10 prostate tissue enriched transcripts by the Human Protein Atlas. FOLH1_HUMAN, also known as prostate-specific membrane antigen (PSMA), is a cell surface protein with enhanced prostate specificity and over-expression in PCa. FOLH1 is currently one of the principal targets of novel PET imaging approaches and radioligand therapies^57^. The prostate-specific protein REL1_HUMAN has never been identified at the protein level in serum, prostate cell lines, and seminal plasma, as we recently demonstrated by immunoassays and highly sensitive targeted IA-PRM assays^58^. Recombinant REL1, however, was efficiently coated onto microplates, as we confirmed with IA-shotgun MS. As a result, we replaced PBS with REL1 protein as an improved negative control for non-specific binding in IA-SRM assays.

Autoantibodies against rKLK4, rPPAP, rFOLH1, and rREL1 were measured by indirect ELISA (**Figure 3A**) and IA-SRM assays (**Figure 3B**). While indirect ELISA could distinguish between a few of the most intense samples as opposed to REL1 negative controls (**Figure 3C**), it revealed the overall high background levels and failed in distinguishing antigen-specific IgG autoantibodies from the matched negative controls. On the contrary, IA-SRM assays revealed much lower background levels in the REL1 negative control group, and enabled detection of much lower levels of rKLK4-, rFOLH1-, or rPPAP-specific IgG1 autoantibodies (**Figures 3B, 3D)**. The limit of detection (LOD) for our IA-SRM assay for IgG1 was estimated at 1 ng/mL (**Supplementary Figure S2**). IgG1 levels were above LOD in all serum samples measured with IA-SRM, including REL1 negative controls (**Figure 3B**).

**Figure 3.**
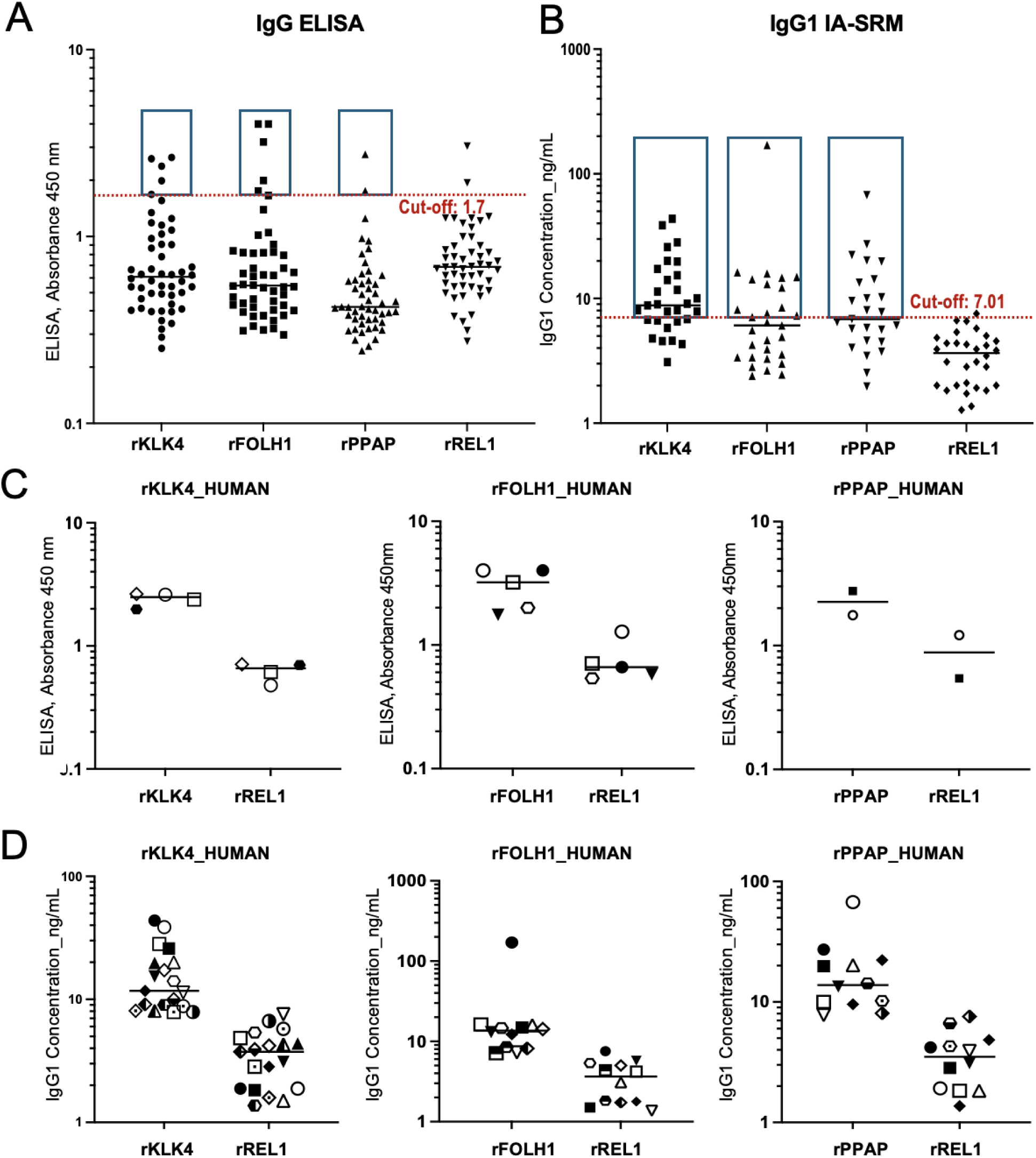
Discovery of serum autoantibodies against prostate-specific antigens rKLK4, rFOLH1, and rPPAP. (**A**) Measurement of serum autoantibodies with indirect ELISA. rREL1 protein was used as a negative control, and immunopositivity was calculated as a mean plus two standard deviations for the rREL1 measurements. Immunopositive samples were highlighted in boxes with the cut-off of 1.3. (**B**) Measurement of serum autoantibodies with IA-SRM. Immunopositive samples were highlighted in boxes with the cut-off of 7.01 ng/mL. (**C, D**) Immunopositive samples matched their negative controls.

In addition to IgG1, our IA-SRM assay detected anti-rKLK3 IgG1, IgA1, and IgM autoantibodies (**Figure 4A**). Raw IA-SRM data for the two representative PCa serum samples with the highest levels of autoantibodies demonstrated 8.8-fold higher levels of anti-rKLK3 IgG1 versus background levels in a matched (-) PBS negative control (**Figure 4B**), as well as 5.8-fold higher levels of anti-rKLK4 IgG1 in a matched (-) REL1 negative control (**Figure 4C**). Raw IA-SRM data comparing independent positive and negative control samples are presented in **Supplementary Figure S3**.

**Figure 4.**
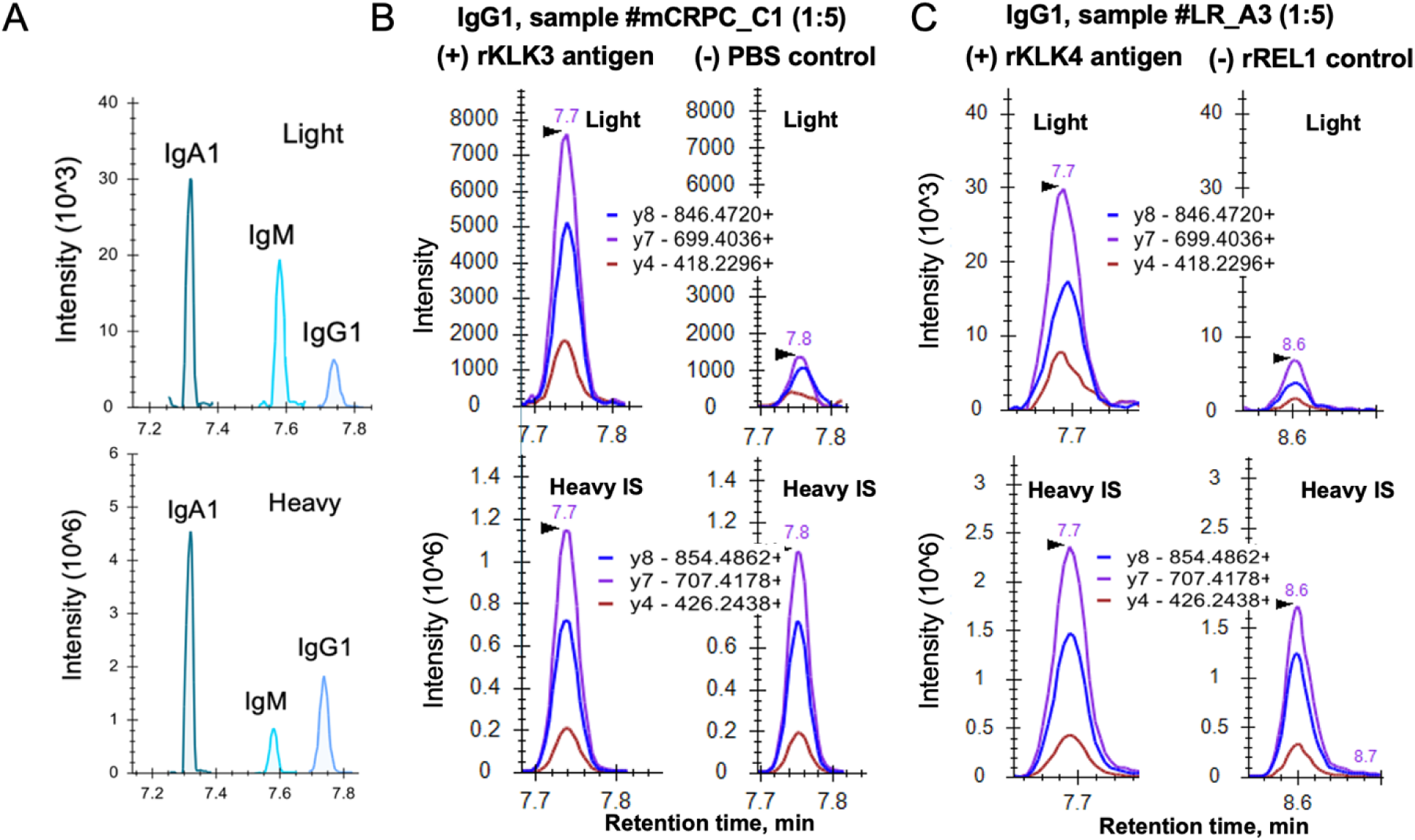
IA-SRM measurements of two representative PCa serum samples with the highest levels of autoantibodies. (**A**) Quantification of anti-rKLK3 IgG1 (peptide GPSVFPLAPSS**K^2+^)**, IgA1 (TPLTATLS**K^2+^)**, and IgM (DGFFGNP**R^2+^**) autoantibodies in a metastatic CRPC sample metC1. (**B**) Raw data for metC1 serum sample demonstrates 8.8-fold higher levels of anti-rKLK3 IgG1 versus background levels in a matched (-) PBS negative control. (**C**) Raw data for low-risk A3 serum sample demonstrates 5.8-fold higher levels of anti-rKLK4 IgG1 versus background levels in a matched (-) rREL1 negative control.

The substantially lower levels of background noise in IA-SRM assays suggested the relatively high levels of non-specific binding in indirect ELISA and could explain poor reproducibility of previous studies on PCa autoantibodies and low immunopositivity in PCa (with a cut-off defined as a mean plus two standard deviations in negative controls).

IA-SRM assays demonstrated the detectable levels of anti-IgG1 autoantibodies against rKLK4, rFOLH1, and rPPAP, with concentrations ranging from ∼8 ng/mL to 170 ng/mL, and the significant differences of their median levels of 8.1 ng/mL (MWU *P*=0.000083), 6.1 ng/mL (*P*=0.0004), and 6.8 ng/mL (P=0.000095), respectively. Interestingly, rKLK4, rFOLH1, and rPPAP have also revealed high levels of IgA1 and IgM autoantibodies (**Supplementary Figure S4**). Our study suggested that IA-SRM was a practical assay to detect and quantify PCa autoantibodies, while indirect ELISA should not be recommended for autoantibody measurements due to its high background.

### Confirmation of autoantibody identity by IA-shotgun MS

To independently confirm autoantibody subtypes, we selected a set of serum samples with th highest concentrations measured by IA-SRM, and enriched autoantibodies with rKLK3, rKLK4, rPPAP, and rFOLH1 antigens, as well as a rREL1 negative control. Following addition of internal standard peptides, digestion, LC-shotgun MS/MS identification, and MSFragger (v 4.1) data analysis, we detected the following isotypes and subclasses of human antibodies in antigen-positive samples (excluding REL1 samples): IGHG1 (6 peptides), IGHG2 (2 peptides), IGHG3 (3 peptides), IGHM (10 peptides), and IGHA1 (6 peptides). Unique IGHG2 and IGHG3 peptides had low intensity, were present in only a few samples, and were excluded from further analysis. Following that, Skyline was used to extract MS1 data for the light endogenous and heavy standard proteotypic peptides of IgG1, IgA1, and IgM (precursor, M+1, M+2), and concentrations were calculated. Median concentrations (**Supplementary Figure S5**) confirmed elevated levels of IgG1, IgA1, and IgM across all antigens. As a result, the phenomenon of autoantibodies in PCa was independently confirmed by IA-SRM and IA-shotgun MS assays. We also confirmed that IgG1 autoantibodies represented a secreted isoform (**Supplementary Figure S6**).

While IgG^52–54^ and IgM^42^ were previously reported as anti-PSA autoantibodies, for the first time, we revealed IgG1, IgM and IgA1 as the principal isotypes of serum autoantibodies against four prostate-specific antigens (**Supplementary Figure S4**).

### Correlation between Indirect ELISA and IA-SRM Assays

To compare the results of indirect ELISA to IA-SRM assays for rKLK3, rKLK4, rFOLH1, and rPPAP, we calculated the immunopositivity cut-offs as a control group mean + 2*SD. Indirect ELISA cut-off was 1.3 (OD 450) for rKLK3 and 1.7 (OD450) for other antigens. IA-SRM cutoffs were 2.8 ng/mL for rKLK3 and 7.01 ng/mL for other antigens (**Figure 5**). Overall, IA-SRM revealed higher immunopositivity, with the majority of PCa samples having the detectable IgG1 levels. Using IA-SRM as a reference assay (with autoantibody positivity confirmed by the presence of LC-SRM peaks), indirect ELISA demonstrated the false negative rates (FNR) between 69 and 92% (**Figure 5**). High FNR of indirect ELSIA and high immunopositivity in IA-SRM assays are explained by the substantially lower background noise of IA-SRM relative to immunoassays.

**Figure 5.**
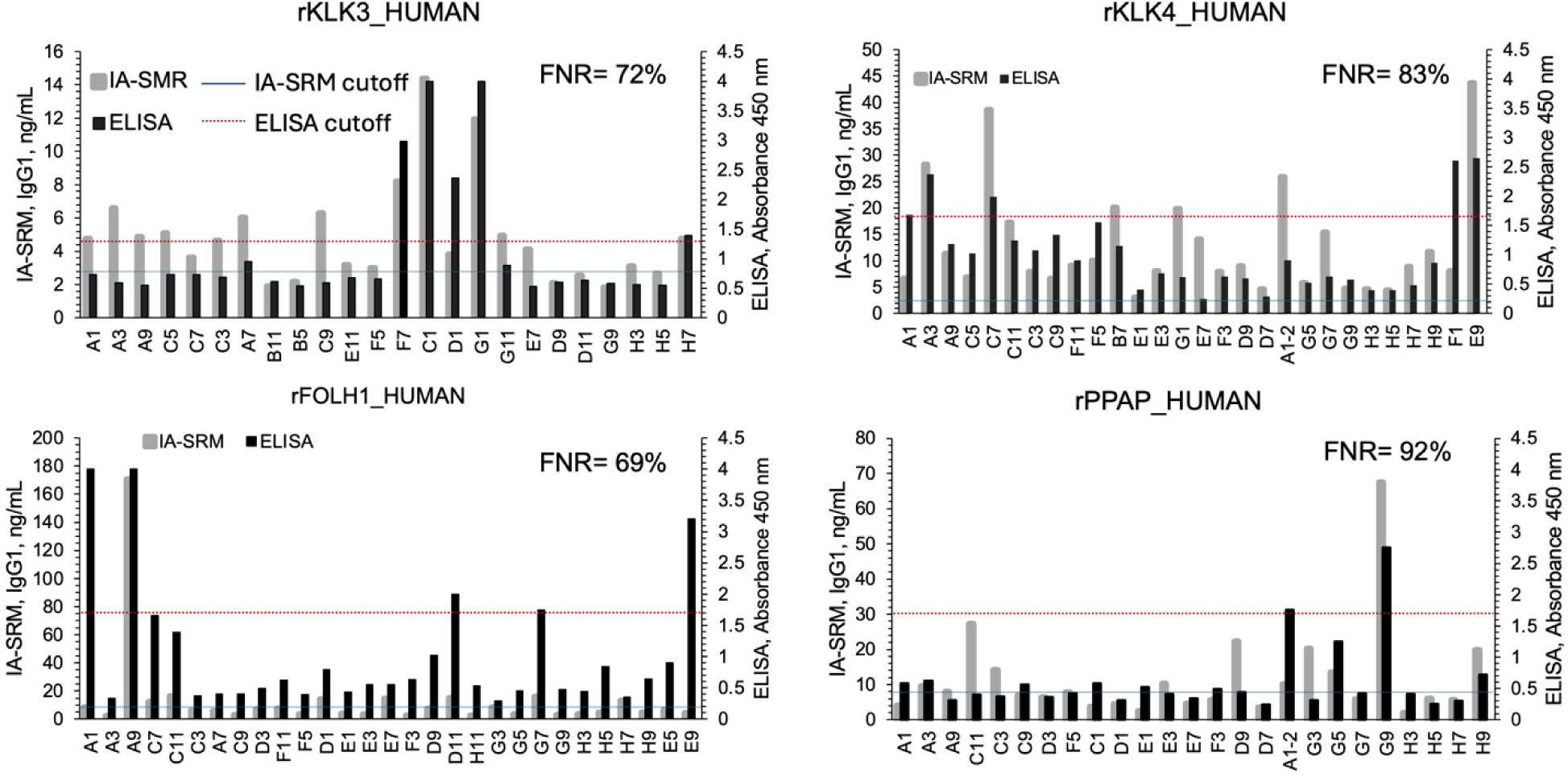
Correlation between ELISA and IA-SRM assays to detect IgG1 autoantibodies against the prostate-specific antigens. Based on their corresponding immunopositivity cutoffs, IA-SRM assays could detect more autoantibody-positive samples than indirect ELISA. Indirect ELISA revealed high false negative rates (FNR) relative to IA-SRM assay.

### Diagnostic performance of autoantibodies

To calculate diagnostic sensitivity and specificity, we used immunopositivity cut-offs calculated as a mean in a control group (PBS or rREL1) plus 2 standard deviations (**Table 2**). PSA (rKLK3) revealed higher sensitivity for IgA1 autoantibodies (75%) compared to IgG1 (66%) and IgM (∼28%). rKLK4 revealed higher sensitivity for IgG1 autoantibodies (66%) compared to IgA1(23%) and IgM (38%). However, AUC for anti-rKLK3 and anti-rKLK4 were the highest for IgG1 (0.69 and 0.66, respectively) compared to IgA1 and IgM (**Table 2**). In addition, neither IgG ELISA nor IgG1 IA-SRM revealed significant diagnostic performance (PCa vs. nBx; 2-tailed Welch’s t-test *P* > 0.05) of anti-rKLK4, -rFOLH1, and -rPPAP autoantibodies (**Figure 6**).

**Figure 6.**
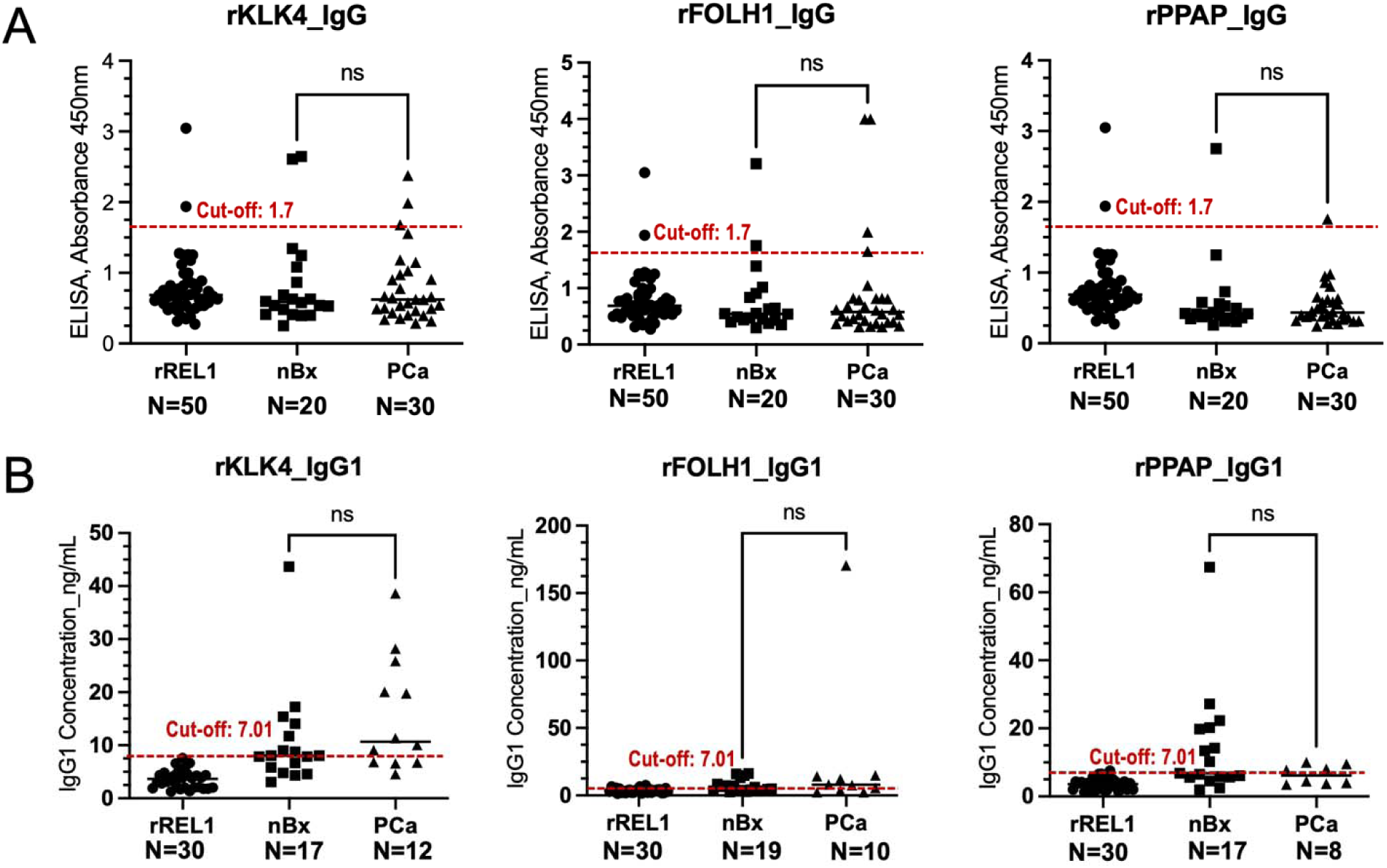
Performance of autoantibodies against rKLK4, rFOLH1, and rPPAP within individual PCa risk groups. (**A**) Antigen-specific IgG measured by indirect ELISA and (**B**) antigen-specific IgG1 measured by IA-SRM assay. Recombinant REL1 was used as a negative control. Neither IgG ELISA nor IgG1 IA-SRM revealed significant diagnostic performances (PCa vs. nBx; 2-tailed Welch’s t-test *P* > 0.05) of anti-rKLK4, -rFOLH1, or -rPPAP autoantibodies. The red dotted line represents cut-off. NS, non-statistically significant.

**Table 2.** AUC and diagnostic sensitivity and specificity of autoantibodies to differentiate between PCa (all groups) and nBx.

|  | PSA (rKLK3) |  |  | rKLK4 |  |  |
| --- | --- | --- | --- | --- | --- | --- |
|  | AUC [CI 95%] | Sensitivity | Specificity | AUC [CI 95%] | Sensitivity | Specificity |
| **IgG1** | 0.69<br>[0.48-0.91] | 66% | 50% | 0.66<br>[0.45-0.87] | 66% | 35% |
| **IgA1** | 0.58<br>[0.34-0.83] | 75% | 25% | 0.61<br>[0.40-0.82] | 23% | 86% |
| **IgM** | 0.66<br>[0.42-0.89] | 29% | 75% | 0.54<br>[0.30-0.77] | 38% | 88% |

## DISCUSSION

PSA (KLK3) is the current biomarker for PCa diagnosis, with serum levels >3 ng/mL considered abnormal^59^. Even though the KLK3 expression levels in prostate cancer cells are decreasing with PCa progression^60^, the disruption of a prostate-blood barrier during prostate enlargement, inflammation, and PCa progression results in leakage of PSA in blood serum. Since PSA is a highly prostate tissue-specific protein, its elevated levels in serum suggest prostate pathologies. Elevated levels of PSA, however, cannot distinguish between PCa and non-malignant abnormalities (enlargement or inflammation), and cannot stratify the PCa risk groups (low-risk, high-risk, and metastatic cancers). While PSA is used clinically, its limitations (low diagnostic specificity and no prognostic value) have spurred the search for novel PCa biomarkers.

Improvements in mass spectrometry instrumentation, robustness of nanoflow and microflow chromatography, and standardization of proteomic sample preparation protocols enabled numerous applications of proteomics in basic research^61–68^ and translational studies^69–80^. We previously evaluated by mass spectrometry and immunoassays numerous prostate-specific proteins including KLK4^32^, TGM4^33^, and REL1^34^. None of these proteins, or the previously evaluated PPAP, outperformed PSA. As an alternative, we aimed here at identification and quantification of the human endogenous autoantibodies against prostate-specific proteins. Our recently developed IA-MS assays to evaluate serological response of infectious diseases^44,46^ served as a foundation to establish IA-MS assays to discover autoantibodies.

Autoantibodies against tumor-associated antigens (TAAs) have gained considerable attention as potential biomarkers for cancer diagnosis^36,37^. Autoantibodies are produced in response to aberrant expression or leakage of tumor-specific and tissue-specific proteins, with an epididymis-specific protein HE4 and its autoantibodies being a prominent example of an ovarian cancer biomarker^81^. In PCa research, numerous studies including top-impact publications^82–86^ claimed diagnostic value of autoantibodies. Autoantibodies were detected for PSA^54^, KLK4^17^, PPAP^52^, and AMACR^87^, even though at low frequency (<10% of patients). The large-scale microarray studies have also demonstrated autoantibodies against non-tissue specific proteins^82,85,88,89^, but failed subsequent validation. In metastatic CRPC, autoantibodies were identified for cancer/testis antigens including CTAG1B (NY-ESO-1)^90,91^, HERVK-113^83^, and AKAP4^92^.

In this study, our central hypothesis was based on previous evidence for prostate being an immune-privileged tissue^93^. Similar to the brain-blood barrier, the prostate-blood barrier excludes prostate-specific proteins from the systemic circulation and systemic immune response. Disruption of the prostate-blood barrier during tumor progression results in leakage of prostate-specific proteins into blood serum, with inducing immune response and producing autoantibodies^36^.

In this discovery study, we utilized IA-SRM assays as novel tools to address the following questions:

i. Are IA-SRM assays more selective than indirect ELISA and have sufficient sensitivity to detect the endogenous human autoantibodies in serum?
ii. Can we provide independent evidence on the presence of autoantibodies against prostate-specific proteins in serum?
iii. Is there a correlation between serum levels of PSA and anti-PSA autoantibodies?
iv. Are there any implications of our findings for the established diagnostics of PCa (assuming serum anti-PSA autoantibody levels of ∼7 ng/mL)?
v. Can we distinguish between PCa and nBx based on autoantibody profiles?
vi. What prostate-specific antigens should be prioritized in future studies?

Conventional assays to detect autoantibodies often rely on indirect immunoassays targeting a single antigen (microplate-based indirect ELISA) or numerous antigens (high-throughput antigen microarrays), with secondary antibodies utilized for detection of primary autoantibodies. Indirect immunoassays provide only semi-quantitative measurements and suffer from cross-reactivity and inability to differentiate between immunoglobulin isotypes and subclasses. Recently, a sensitive and multiplex but indirect ADAP assay (Antibody detection by agglutination-PCR) was demonstrated as a novel method to measure multiple antibodies/autoantibodies pairs^94^. Autoantibodies, however, have never been validated using direct assays such as IA-SRM. As a result, there was certain skepticism regarding the evidence for the endogenous autoantibodies against prostate-specific antigens and their value for PCa diagnostics.

In this study, we demonstrated that IA-SRM was superior to indirect ELISA due to the high analytical selectivity and sufficient sensitivity. Indirect ELISA revealed a high background that resulted in a high rate of false negative identifications. We evaluated by indirect ELISA and IA-SRM the autoantibodies against several prostate-specific proteins including PSA, KLK4, PPAP, FOLH1, and REL1. The recombinant REL1 was utilized as a negative control since the endogenous REL1 transcript was not expressed at the protein level^34^, as we recently demonstrated.

Here, we detected anti-IgG1 autoantibodies against PSA (61% of all samples), rKLK4 (66%), rFOLH1 (45%), and rPPAP (48%) antigens. Anti-rKLK4 autoantibodies were detected at higher concentrations than anti-rKLK3 autoantibodies, making rKLK4 a promising candidate for further investigation in larger cohorts. IA-shotgun MS/MS provided further evidence on the identity of antibody isotypes and subclasses, with IgG1, IgA1 and IgM being the most abundant isotypes.

Due to the lack of reference standards of the human endogenous polyclonal antibodies against each antigen, previous immunoassay studies calibrated the autoantibody-mediated absorption to the dilution series of the total human polyclonal IgG. Surprisingly, such calibration resulted in 1,000-fold over-estimated levels of anti-PSA antibodies in the range of µg/mL, such as 5.7 µg/mL^55^. Furthermore, the reported concentrations of polyclonal IgG autoantibodies exceeded the levels of circulating PSA by 200-500 fold ^41,55,95^. If true, such enormous excess of serum polyclonal autoantibodies would completely saturate serum PSA (assuming polyclonal autoantibodies mask all epitopes) and make the detection of circulating PSA impossible if using sandwich immunoassays. In other words, the serum PSA-mediated PCa diagnostics would not work. Most probably, the high autoantibody levels reported in some previous studies were due to lack of reference standards and non-specific binding of indirect immunoassays. Here, we detected anti-IgG1 autoantibodies against PSA, rKLK4, rFOLH1, and rPPAP antigens present at substantially lower levels of ng/mL levels (1.9 ng/mL to 170 ng/mL). Assuming high affinity 1:1 binding, the median 5 ng/mL (34 fmol/mL) of IgG1 would bind only 1.1 ng/mL (34 fmol/mL) of circulating PSA. In this case, the measured PSA values (a clinical cutoff of >4 ng/mL or 121 fmol/mL) would be underestimated by only ∼1.1 ng/mL. These estimates highlight the importance of independent IA-SRM assays for quantification of autoantibodies and resolving the controversies on their over-estimated µg/mL levels.

We demonstrated that anti-rKLK3 autoantibodies could differentiate nBx from PCa. While autoantibodies may not serve as standalone biomarkers for PCa diagnosis, their measurement in serum could provide valuable insights into a patient’s immune response to prostate-specific antigens. Previous studies have shown that combinations of PSA levels with autoantibody measurements could increase diagnostic specificity to distinguish PCa from benign prostatic hyperplasia (defined here as a negative biopsy group)^96^. Consequently, integrating autoantibody detection alongside PSA measurement could expand the scope of PCa diagnosis and improve patient identification, particularly in cases where PSA alone may be insufficient. Our study revealed that PSA, rKLK4 and potentially rPPAP could be prioritized in future studies on measuring autoantibodies in large cohorts of PCa serum samples.

In future studies, our IA-SRM assays may emerge as a complimentary diagnostic tool to improve PCa immunotherapy through detecting pre-existing autoantibodies and monitoring therapy-induced autoantibodies^97^. Currently, prostate (PSA^98^, KLK4^17^, FOLH1^99,100^, PPAP^101,102^, STEAP1^103^) and testis (CTAG1B^91^, MAGEA2^104^, AKAP4^92^) antigens are being evaluated as innovative immunotherapy targets to treat deadly mCRPC. Pre-existing autoantibodies against tumor-associated antigens can impair the efficacy of immune-based treatments and immune checkpoint inhibitors by inducing immune exhaustion and preventing sufficient immune activation ^105–107^. For example, patients with the high pre-existing levels of autoantibodies against PPAP may not respond well to PPAP-targeting immunotherapy^101,102^ and may need to be treated with alternatives therapies targeting PSA^98^ or FOLH1^99,100^.

The knowledge of the autoantibody levels may also improve the emerging molecular imaging diagnostics such as positron emission tomography (PET) approaches to detect metastatic mCRPC^58^. FOLH1 (PSMA) is a widely used target for prostate cancer imaging using radiolabeled anti-FOLH1 monoclonal antibodies^108, 109^. Pre-existing autoantibodies (for example, 170 ng/mL anti-FOLH1 autoantibodies detected in our study) could compete with the molecular imaging agent and mask PET detection of metastatic lesions. Such challenges were recognized for molecular imaging of thyroid cancer and emphasizing the need for pre-screening strategies for antithyroglobulin antibodies^110^.

## CONCLUSIONS

In conclusion, our study provided evidence for autoantibodies against prostate-specific antigens, with IA-SRM assays offering a more specific alternative to ELISA. We demonstrated that PSA autoantibodies have potential diagnostic value in differentiating nBx from PCa samples. While autoantibodies may not serve as standalone diagnostic biomarkers for PCa, their integration with diagnostic and therapeutic approaches may improve stratification of patients for immunotherapies and molecular diagnostic imaging. IA-SRM assays may facilitate discovery of novel autoantibody targets and elucidate the mechanisms of immune response and immunotherapy resistance. Ultimately, the IA-SRM assays offer a novel platform for autoimmune diagnostics and cancer diagnostics, paving the way for more personalized and precise approaches to cancer detection and management.

## Data Availability

All data produced in the present study are available upon reasonable request to the authors.

## Notes

The authors declare no competing financial interest.

## Funding

Cancer Research Society 2024 CRS Operating Grant (#1281719) and Canadian Institutes of Health Research-Institute of Cancer Research / Cancer Research Society Partnership - Research Grant (#196966) to APD.

## ACKNOWLEDGMENTS

We thank the Alberta Proteomics and Mass Spectrometry Facility and Jack Moore for assistance with mass spectrometry instruments, and Alberta Prostate Cancer Registry & Biorepository for providing blood serum samples.

## ABBREVIATIONS

ELISA: Enzyme-linked immunosorbent assay
IA-MS: Immunoaffinity-mass spectrometry
LC-MS: liquid chromatography–mass spectrometry
LOD: Limit of detection
ppm: parts per million
SRM: Selected reaction monitoring
mCRPC: metastatic castration-resistant prostate cancer
PCa: Prostate Cancer
KLK3: Kallikrein 3
KLK4: Kallikrein 4
FOLH1: Folate Hydrolase 1
PPAP: Prostatic acid phosphatase
REL1: Relaxin 1
NS: non-statistically significant.

## Notes

### Competing Interest Statement

The authors have declared no competing interest.

### Author Declarations

Cancer Committee of Health Research Ethics Board of Alberta (HREBA) gave ethical approval for this work (HREBA.CC-22-0056 and HREBA.CC-14-0085).

## REFERENCES

1. 2021, C. c. s. Canadian Cancer Society 2021, https://cdn.cancer.ca/-/media/files/research/cancer-statistics/2021-statistics/2021-pdf-en-final.pdf.

(2) Barry, M. J. Clinical practice. Prostate-specific-antigen testing for early diagnosis of prostate cancer. N Engl J Med 2001, 344, 1373–1377, 10.1056/NEJM200105033441806

(3) Collin, S. M.; Martin, R. M.; Metcalfe, C.; Gunnell, D.; Albertsen, P. C.; Neal, D.; Hamdy, F.; Stephens, P.; Lane, J. A.; Moore, R.; Donovan, J. Prostate-cancer mortality in the USA and UK in 1975-2004: an ecological study. Lancet Oncol 2008, 9, 445–452, 10.1016/S1470-2045(08)70104-9

(4) Heijnsdijk, E. A.; Wever, E. M.; Auvinen, A.; Hugosson, J.; Ciatto, S.; Nelen, V.; Kwiatkowski, M.; Villers, A.; Paez, A.; Moss, S. M.; Zappa, M.; Tammela, T. L. et al. Quality-of-life effects of prostate-specific antigen screening. N Engl J Med 2012, 367, 595–605, 10.1056/NEJMoa1201637

(5) Mason, R. J.; Marzouk, K.; Finelli, A.; Saad, F.; So, A. I.; Violette, P. D.; Breau, R. H.; Rendon, R. A. UPDATE - 2022 Canadian Urological Association recommendations on prostate cancer screening and early diagnosis Endorsement of the 2021 Cancer Care Ontario guidelines on prostate multiparametric magnetic resonance imaging - Summary of changes. Can Urol Assoc J 2022, 16, 86–87, 10.5489/cuaj.7832

(6) Schroder, F. H.; Hugosson, J.; Roobol, M. J.; Tammela, T. L.; Ciatto, S.; Nelen, V.; Kwiatkowski, M.; Lujan, M.; Lilja, H.; Zappa, M.; Denis, L. J.; Recker, F. et al. Prostate-cancer mortality at 11 years of follow-up. N Engl J Med 2012, 366, 981–990, 10.1056/NEJMoa1113135

(7) Wilt, T. J.; Brawer, M. K.; Jones, K. M.; Barry, M. J.; Aronson, W. J.; Fox, S.; Gingrich, J. R.; Wei, J. T.; Gilhooly, P.; Grob, B. M.; Nsouli, I.; Iyer, P. et al. Radical prostatectomy versus observation for localized prostate cancer. N Engl J Med 2012, 367, 203–213, 10.1056/NEJMoa1113162

(8) Sandhu, S.; Moore, C. M.; Chiong, E.; Beltran, H.; Bristow, R. G.; Williams, S. G. Prostate cancer. Lancet 2021, 398, 1075–1090, 10.1016/S0140-6736(21)00950-8

(9) Sridaran, D.; Bradshaw, E.; DeSelm, C.; Pachynski, R.; Mahajan, K.; Mahajan, N. P. Prostate cancer immunotherapy: Improving clinical outcomes with a multi-pronged approach. Cell Rep Med 2023, 4, 101199, 10.1016/j.xcrm.2023.101199

(10) Zhang, Y.; Zhang, Z. The history and advances in cancer immunotherapy: understanding the characteristics of tumor-infiltrating immune cells and their therapeutic implications. Cell Mol Immunol 2020, 17, 807–821, 10.1038/s41423-020-0488-6

(11) Beer, T. M.; Kwon, E. D.; Drake, C. G.; Fizazi, K.; Logothetis, C.; Gravis, G.; Ganju, V.; Polikoff, J.; Saad, F.; Humanski, P.; Piulats, J. M.; Gonzalez Mella, P. et al. Randomized, Double-Blind, Phase III Trial of Ipilimumab Versus Placebo in Asymptomatic or Minimally Symptomatic Patients With Metastatic Chemotherapy-Naive Castration-Resistant Prostate Cancer. J Clin Oncol 2017, 35, 40–47, 10.1200/JCO.2016.69.1584

(12) Kwon, E. D.; Drake, C. G.; Scher, H. I.; Fizazi, K.; Bossi, A.; van den Eertwegh, A. J.; Krainer, M.; Houede, N.; Santos, R.; Mahammedi, H.; Ng, S.; Maio, M., et al. Ipilimumab versus placebo after radiotherapy in patients with metastatic castration-resistant prostate cancer that had progressed after docetaxel chemotherapy (CA184-043): a multicentre, randomised, double-blind, phase 3 trial. Lancet Oncol 2014, 15, 700–712, 10.1016/S1470-2045(14)70189-5

(13) Lu, X.; Horner, J. W.; Paul, E.; Shang, X.; Troncoso, P.; Deng, P.; Jiang, S.; Chang, Q.; Spring, D. J.; Sharma, P.; Zebala, J. A.; Maeda, D. Y. et al. Effective combinatorial immunotherapy for castration-resistant prostate cancer. Nature 2017, 543, 728–732, 10.1038/nature21676

(14) Rastogi, I.; Muralidhar, A.; McNeel, D. G. Vaccines as treatments for prostate cancer. Nat Rev Urol 2023, 20, 544–559, 10.1038/s41585-023-00739-w

(15) Shore, N. D.; Morrow, M. P.; McMullan, T.; Kraynyak, K. A.; Sylvester, A.; Bhatt, K.; Cheung, J.; Boyer, J. D.; Liu, L.; Sacchetta, B.; Rosencranz, S.; Heath, E. I. et al. CD8(+) T Cells Impact Rising PSA in Biochemically Relapsed Cancer Patients Using Immunotherapy Targeting Tumor-Associated Antigens. Mol Ther 2020, 28, 1238–1250, 10.1016/j.ymthe.2020.02.018

(16) Lopez-Bujanda, Z. A.; Obradovic, A.; Nirschl, T. R.; Crowley, L.; Macedo, R.; Papachristodoulou, A.; O’Donnell, T.; Laserson, U.; Zarif, J. C.; Reshef, R.; Yuan, T.; Soni, M. K. et al. TGM4: an immunogenic prostate-restricted antigen. J Immunother Cancer 2021, *9*, 10.1136/jitc-2020-001649

(17) Wilkinson, R.; Woods, K.; D’Rozario, R.; Prue, R.; Vari, F.; Hardy, M. Y.; Dong, Y.; Clements, J. A.; Hart, D. N. J.; Radford, K. J. Human kallikrein 4 signal peptide induces cytotoxic T cell responses in healthy donors and prostate cancer patients. Cancer Immunol Immunother 2012, 61, 169–179, 10.1007/s00262-011-1095-2

(18) Drabovich, A. P.; Saraon, P.; Jarvi, K.; Diamandis, E. P. Seminal plasma as a diagnostic fluid for male reproductive system disorders. Nat Rev Urol 2014, 11, 278–288, 10.1038/nrurol.2014.74

(19) Cha, H. R.; Lee, J. H.; Ponnazhagan, S. Revisiting Immunotherapy: A Focus on Prostate Cancer. Cancer Res 2020, 80, 1615–1623, 10.1158/0008-5472.CAN-19-2948

(20) Al Olama, A. A.; Kote-Jarai, Z.; Giles, G. G.; Guy, M.; Morrison, J.; Severi, G.; Leongamornlert, D. A.; Tymrakiewicz, M.; Jhavar, S.; Saunders, E.; Hopper, J. L.; Southey, M. C. et al. Multiple loci on 8q24 associated with prostate cancer susceptibility. Nat Genet 2009, 41, 1058–1060

(21) Eeles, R. A.; Kote-Jarai, Z.; Al Olama, A. A.; Giles, G. G.; Guy, M.; Severi, G.; Muir, K.; Hopper, J. L.; Henderson, B. E.; Haiman, C. A.; Schleutker, J.; Hamdy, F. C. et al. Identification of seven new prostate cancer susceptibility loci through a genome-wide association study. Nat Genet 2009, 41, 1116–1121

(22) Gudmundsson, J.; Sulem, P.; Gudbjartsson, D. F.; Blondal, T.; Gylfason, A.; Agnarsson, B. A.; Benediktsdottir, K. R.; Magnusdottir, D. N.; Orlygsdottir, G.; Jakobsdottir, M.; Stacey, S. N.; Sigurdsson, A. et al. Genome-wide association and replication studies identify four variants associated with prostate cancer susceptibility. Nat Genet 2009, 41, 1122–1126, 10.1038/ng.448

(23) Haiman, C. A.; Patterson, N.; Freedman, M. L.; Myers, S. R.; Pike, M. C.; Waliszewska, A.; Neubauer, J.; Tandon, A.; Schirmer, C.; McDonald, G. J.; Greenway, S. C.; Stram, D. O. et al. Multiple regions within 8q24 independently affect risk for prostate cancer. Nat Genet 2007, 39, 638–644, 10.1038/ng2015

(24) Ahn, J.; Berndt, S. I.; Wacholder, S.; Kraft, P.; Kibel, A. S.; Yeager, M.; Albanes, D.; Giovannucci, E.; Stampfer, M. J.; Virtamo, J.; Thun, M. J.; Feigelson, H. S. et al. Variation in KLK genes, prostate-specific antigen and risk of prostate cancer. Nat Genet 2008, 40, 1032–1034; author reply 1035-1036, 10.1038/ng0908-1032

(25) Farha, M. W.; Salami, S. S. Biomarkers for prostate cancer detection and risk stratification. Ther Adv Urol 2022, 14, 17562872221103988, 10.1177/17562872221103988

(26) Bussemakers, M. J.; van Bokhoven, A.; Verhaegh, G. W.; Smit, F. P.; Karthaus, H. F.; Schalken, J. A.; Debruyne, F. M.; Ru, N.; Isaacs, W. B. DD3: a new prostate-specific gene, highly overexpressed in prostate cancer. Cancer Res 1999, 59, 5975–5979

(27) Day, J. R.; Jost, M.; Reynolds, M. A.; Groskopf, J.; Rittenhouse, H. PCA3: from basic molecular science to the clinical lab. Cancer Lett 2011, 301, 1–6, 10.1016/j.canlet.2010.10.019

(28) Hessels, D.; Smit, F. P.; Verhaegh, G. W.; Witjes, J. A.; Cornel, E. B.; Schalken, J. A. Detection of TMPRSS2-ERG fusion transcripts and prostate cancer antigen 3 in urinary sediments may improve diagnosis of prostate cancer. Clin Cancer Res 2007, 13, 5103–5108, 10.1158/1078-0432.CCR-07-0700

(29) Khoo, A.; Govindarajan, M.; Qiu, Z.; Liu, L. Y.; Ignatchenko, V.; Waas, M.; Macklin, A.; Keszei, A.; Neu, S.; Main, B. P.; Yang, L.; Lance, R. S. et al. Prostate cancer reshapes the secreted and extracellular vesicle urinary proteomes. Nat Commun 2024, 15, 5069, 10.1038/s41467-024-49424-5

(30) Kim, Y.; Ignatchenko, V.; Yao, C. Q.; Kalatskaya, I.; Nyalwidhe, J. O.; Lance, R. S.; Gramolini, A. O.; Troyer, D. A.; Stein, L. D.; Boutros, P. C.; Medin, J. A.; Semmes, O. J. et al. Identification of differentially expressed proteins in direct expressed prostatic secretions of men with organ-confined versus extracapsular prostate cancer. Mol Cell Proteomics 2012, 11, 1870–1884, 10.1074/mcp.M112.017889

(31) Kim, Y.; Jeon, J.; Mejia, S.; Yao, C. Q.; Ignatchenko, V.; Nyalwidhe, J. O.; Gramolini, A. O.; Lance, R. S.; Troyer, D. A.; Drake, R. R.; Boutros, P. C.; Semmes, O. J. et al. Targeted proteomics identifies liquid-biopsy signatures for extracapsular prostate cancer. Nat Commun 2016, 7, 11906, 10.1038/ncomms11906

(32) Karakosta, T. D.; Soosaipillai, A.; Diamandis, E. P.; Batruch, I.; Drabovich, A. P. Quantification of Human Kallikrein-Related Peptidases in Biological Fluids by Multiplatform Targeted Mass Spectrometry Assays. Mol Cell Proteomics 2016, 15, 2863–2876, 10.1074/mcp.M115.057695

(33) Drabovich, A. P.; Saraon, P.; Drabovich, M.; Karakosta, T. D.; Dimitromanolakis, A.; Hyndman, M. E.; Jarvi, K.; Diamandis, E. P. Multi-omics biomarker pipeline reveals elevated levels of Protein-glutamine Gamma-glutamyltransferase 4 in seminal plasma of prostate cancer patients. Mol Cell Proteomics 2019, 18, 1807–1823, 10.1074/mcp.RA119.001612

(34) Rais, Y.; Drabovich, A. P. Identification and Quantification of Human Relaxin Proteins by Immunoaffinity-Mass Spectrometry. J Proteome Res 2024, 23, 2013–2027, 10.1021/acs.jproteome.4c00027

(35) Fu, Z.; Rais, Y.; Bismar, T. A.; Hyndman, M. E.; Le, X. C.; Drabovich, A. P. Mapping isoform abundance and interactome of the endogenous TMPRSS2-ERG fusion protein by orthogonal immunoprecipitation-mass spectrometry assays. Mol Cell Proteomics 2021, 100075, 10.1016/j.mcpro.2021.100075

(36) Zaenker, P.; Gray, E. S.; Ziman, M. R. Autoantibody Production in Cancer--The Humoral Immune Response toward Autologous Antigens in Cancer Patients. Autoimmun Rev 2016, 15, 477–483, 10.1016/j.autrev.2016.01.017

(37) Young Han, C.; Bedia, J. S.; Yang, W. L.; Hawley, S. J.; Bergan, L.; Hopper, M.; Celestino, J.; Guo, J.; Gornet, T. G.; Soosaipillai, A.; Yang, H.; Doskocil, S. D., et al. Autoantibodies, antigen-autoantibody complexes and antigens complement CA125 for early detection of ovarian cancer. Br J Cancer 2024, 10.1038/s41416-023-02560-z

(38) Henderson, M. C.; Silver, M.; Tran, Q.; Letsios, E. E.; Mulpuri, R.; Reese, D. E.; Lourenco, A. P.; LaBaer, J.; Anderson, K. S.; Alpers, J.; Costantini, C.; Rohatgi, N. et al. A Noninvasive Blood-based Combinatorial Proteomic Biomarker Assay to Detect Breast Cancer in Women over age 50 with BI-RADS 3, 4, or 5 Assessment. Clin Cancer Res 2019, 25, 142–149, 10.1158/1078-0432.CCR-18-0843

(39) Sullivan, F. M.; Mair, F. S.; Anderson, W.; Armory, P.; Briggs, A.; Chew, C.; Dorward, A.; Haughney, J.; Hogarth, F.; Kendrick, D.; Littleford, R.; McConnachie, A. et al. Earlier diagnosis of lung cancer in a randomised trial of an autoantibody blood test followed by imaging. Eur Respir J 2021, *57*, 10.1183/13993003.00670-2020

(40) Moghaddas, F.; Joshua, F.; Taylor, R.; Fritzler, M. J.; Toh, B. H. Autoantibodies directed to centromere protein F in a patient with BRCA1 gene mutation. BMC Res Notes 2016, 9, 84, 10.1186/s13104-016-1908-7

(41) Macdonald, I. K.; Parsy-Kowalska, C. B.; Chapman, C. J. Autoantibodies: Opportunities for Early Cancer Detection. Trends Cancer 2017, 3, 198–213, 10.1016/j.trecan.2017.02.003

(42) Beneduce, L.; Prayer-Galetti, T.; Giustinian, A. M.; Gallotta, A.; Betto, G.; Pagano, F.; Fassina, G. Detection of prostate-specific antigen coupled to immunoglobulin M in prostate cancer patients. Cancer Detect Prev 2007, 31, 402–407, 10.1016/j.cdp.2007.10.005

(43) Lautenbach, N.; Muntener, M.; Zanoni, P.; Saleh, L.; Saba, K.; Umbehr, M.; Velagapudi, S.; Hof, D.; Sulser, T.; Wild, P. J.; von Eckardstein, A.; Poyet, C. Prevalence and causes of abnormal PSA recovery. Clin Chem Lab Med 2018, 56, 341–349, 10.1515/cclm-2017-0246

(44) Fu, Z.; Rais, Y.; Dara, D.; Jackson, D.; Drabovich, A. P. Rational Design and Development of SARS-CoV-2 Serological Diagnostics by Immunoprecipitation-Targeted Proteomics. Anal Chem 2022, 94, 12990–12999, 10.1021/acs.analchem.2c01325

(45) Walter, J.; Eludin, Z.; Drabovich, A. P. Redefining serological diagnostics with immunoaffinity proteomics. Clin Proteomics 2023, 20, 42, 10.1186/s12014-023-09431-y

(46) Tang, W.; Drabovich, A. P. The proteomic toolbox for identification, quantification, and characterization of polyclonal antibodies. bioRxiv 2023, 2023.2010.2027.564451, 10.1101/2023.10.27.564451

(47) Rais, Y.; Fu, Z.; Drabovich, A. P. Mass spectrometry-based proteomics in basic and translational research of SARS-CoV-2 coronavirus and its emerging mutants. Clin Proteomics 2021, 18, 19, 10.1186/s12014-021-09325-x

(48) Tsantilas, K. A.; Merrihew, G. E.; Robbins, J. E.; Johnson, R. S.; Park, J.; Plubell, D. L.; Canterbury, J. D.; Huang, E.; Riffle, M.; Sharma, V.; MacLean, B. X.; Eckels, J. et al. A Framework for Quality Control in Quantitative Proteomics. J Proteome Res 2024, 23, 4392–4408, 10.1021/acs.jproteome.4c00363

(49) Wichmann, C.; Meier, F.; Virreira Winter, S.; Brunner, A. D.; Cox, J.; Mann, M. MaxQuant. Live Enables Global Targeting of More Than 25,000 Peptides. Mol Cell Proteomics 2019, 18, 982–994, 10.1074/mcp.TIR118.001131

(50) Sanchez, T. W.; Zhang, G.; Li, J.; Dai, L.; Mirshahidi, S.; Wall, N. R.; Yates, C.; Wilson, C.; Montgomery, S.; Zhang, J. Y.; Casiano, C. A. Immunoseroproteomic Profiling in African American Men with Prostate Cancer: Evidence for an Autoantibody Response to Glycolysis and Plasminogen-Associated Proteins. Mol Cell Proteomics 2016, 15, 3564–3580, 10.1074/mcp.M116.060244

(51) Gomez-Banuelos, E.; Yu, Y.; Li, J.; Cashman, K. S.; Paz, M.; Trejo-Zambrano, M. I.; Bugrovsky, R.; Wang, Y.; Chida, A. S.; Sherman-Baust, C. A.; Ferris, D. P.; Goldman, D. W. et al. Affinity maturation generates pathogenic antibodies with dual reactivity to DNase1L3 and dsDNA in systemic lupus erythematosus. Nat Commun 2023, 14, 1388, 10.1038/s41467-023-37083-x

(52) Potluri, H. K.; Ng, T. L.; Newton, M. A.; Zhang, J.; Maher, C. A.; Nelson, P. S.; McNeel, D. G. Antibody profiling of patients with prostate cancer reveals differences in antibody signatures among disease stages. J Immunother Cancer 2020, *8*, 10.1136/jitc-2020-001510

(53) Lokant, M. T.; Naz, R. K. Presence of PSA auto-antibodies in men with prostate abnormalities (prostate cancer/benign prostatic hyperplasia/prostatitis). Andrologia 2015, 47, 328–332, 10.1111/and.12265

(54) McNeel, D. G.; Nguyen, L. D.; Storer, B. E.; Vessella, R.; Lange, P. H.; Disis, M. L. Antibody immunity to prostate cancer associated antigens can be detected in the serum of patients with prostate cancer. J Urol 2000, 164, 1825–1829

(55) Nakajima, K.; Heilbrun, L. K.; Smith, D.; Hogan, V.; Raz, A.; Heath, E. The influence of PSA autoantibodies in prostate cancer patients: a prospective clinical study-II. Oncotarget 2017, 8, 17643–17650, 10.18632/oncotarget.12620

(56) Kammeijer, G. S. M.; Nouta, J.; de la Rosette, J.; de Reijke, T. M.; Wuhrer, M. An In-Depth Glycosylation Assay for Urinary Prostate-Specific Antigen. Anal Chem 2018, 90, 4414–4421, 10.1021/acs.analchem.7b04281

(57) Eiber, M.; Fendler, W. P.; Rowe, S. P.; Calais, J.; Hofman, M. S.; Maurer, T.; Schwarzenboeck, S. M.; Kratowchil, C.; Herrmann, K.; Giesel, F. L. Prostate-Specific Membrane Antigen Ligands for Imaging and Therapy. J Nucl Med 2017, 58, 67S–76S, 10.2967/jnumed.116.186767

(58) Aebersold, R.; Agar, J. N.; Amster, I. J.; Baker, M. S.; Bertozzi, C. R.; Boja, E. S.; Costello, C. E.; Cravatt, B. F.; Fenselau, C.; Garcia, B. A.; Ge, Y.; Gunawardena, J. et al. How many human proteoforms are there? Nat Chem Biol 2018, 14, 206–214, 10.1038/nchembio.2576

(59) Atan, A.; Guzel, O. How should prostate specific antigen be interpreted? Turk J Urol 2013, 39, 188–193, 10.5152/tud.2013.038

(60) Tang, D. G. Understanding and targeting prostate cancer cell heterogeneity and plasticity. Semin Cancer Biol 2022, 82, 68–93, 10.1016/j.semcancer.2021.11.001

(61) Fu, Z.; Rais, Y.; Bismar, T. A.; Hyndman, M. E.; Le, X. C.; Drabovich, A. P. Mapping Isoform Abundance and Interactome of the Endogenous TMPRSS2-ERG Fusion Protein by Orthogonal Immunoprecipitation-Mass Spectrometry Assays. Mol Cell Proteomics 2021, 20, 100075, 10.1016/j.mcpro.2021.100075

(62) Schiza, C.; Korbakis, D.; Panteleli, E.; Jarvi, K.; Drabovich, A. P.; Diamandis, E. P. Discovery of a Human Testis-specific Protein Complex TEX101-DPEP3 and Selection of Its Disrupting Antibodies. Mol Cell Proteomics 2018, 17, 2480–2495, 10.1074/mcp.RA118.000749

(63) Korbakis, D.; Brinc, D.; Schiza, C.; Soosaipillai, A.; Jarvi, K.; Drabovich, A. P.; Diamandis, E. P. Immunocapture-Selected Reaction Monitoring Screening Facilitates the Development of ELISA for the Measurement of Native TEX101 in Biological Fluids. Mol Cell Proteomics 2015, 14, 1517–1526, 10.1074/mcp.M114.047571

(64) Drabovich, A. P.; Pavlou, M. P.; Dimitromanolakis, A.; Diamandis, E. P. Quantitative analysis of energy metabolic pathways in MCF-7 breast cancer cells by selected reaction monitoring assay. Mol Cell Proteomics 2012, 11, 422–434, 10.1074/mcp.M111.015214

(65) Begcevic, I.; Brinc, D.; Dukic, L.; Simundic, A. M.; Zavoreo, I.; Basic Kes, V.; Martinez-Morillo, E.; Batruch, I.; Drabovich, A. P.; Diamandis, E. P. Targeted mass spectrometry-based assays for relative quantification of 30 brain-related proteins and their clinical applications. J Proteome Res 2018, 17, 2282–2292, 10.1021/acs.jproteome.7b00768

(66) Martinez-Morillo, E.; Nielsen, H. M.; Batruch, I.; Drabovich, A. P.; Begcevic, I.; Lopez, M. F.; Minthon, L.; Bu, G.; Mattsson, N.; Portelius, E.; Hansson, O.; Diamandis, E. P. Assessment of peptide chemical modifications on the development of an accurate and precise multiplex selected reaction monitoring assay for apolipoprotein e isoforms. J Proteome Res 2014, 13, 1077–1087, 10.1021/pr401060x

(67) Drabovich, A. P.; Pavlou, M. P.; Schiza, C.; Diamandis, E. P. Dynamics of protein expression reveals primary targets and secondary messengers of estrogen receptor alpha signaling in MCF-7 breast cancer cells. Mol Cell Proteomics 2016, 15, 2093–2107, 10.1074/mcp.M115.057257

(68) Dimitrakopoulos, L.; Prassas, I.; Diamandis, E. P.; Nesvizhskii, A.; Kislinger, T.; Jaffe, J.; Drabovich, A. Proteogenomics: Opportunities and Caveats. Clin Chem 2016, 62, 551–557, 10.1373/clinchem.2015.247858

(69) Zhang, J.; Kanoatov, M.; Jarvi, K.; Gauthier-Fisher, A.; Moskovtsev, S. I.; Librach, C.; Drabovich, A. P. Germ cell-specific proteins AKAP4 and ASPX facilitate identification of rare spermatozoa in non-obstructive azoospermia. Mol Cell Proteomics 2023, 22, 100556, 10.1016/j.mcpro.2023.100556

(70) Schiza, C.; Korbakis, D.; Jarvi, K.; Diamandis, E. P.; Drabovich, A. P. Identification of TEX101-associated Proteins Through Proteomic Measurement of Human Spermatozoa Homozygous for the Missense Variant rs35033974. Mol Cell Proteomics 2019, 18, 338–351, 10.1074/mcp.RA118.001170

(71) Korbakis, D.; Schiza, C.; Brinc, D.; Soosaipillai, A.; Karakosta, T. D.; Legare, C.; Sullivan, R.; Mullen, B.; Jarvi, K.; Diamandis, E. P.; Drabovich, A. P. Preclinical evaluation of a TEX101 protein ELISA test for the differential diagnosis of male infertility. BMC Med 2017, 15, 60, 10.1186/s12916-017-0817-5

(72) Drabovich, A. P.; Dimitromanolakis, A.; Saraon, P.; Soosaipillai, A.; Batruch, I.; Mullen, B.; Jarvi, K.; Diamandis, E. P. Differential diagnosis of azoospermia with proteomic biomarkers ECM1 and TEX101 quantified in seminal plasma. Sci Transl Med 2013, 5, 212ra160, 10.1126/scitranslmed.3006260

(73) Cho, C. K.; Drabovich, A. P.; Karagiannis, G. S.; Martinez-Morillo, E.; Dason, S.; Dimitromanolakis, A.; Diamandis, E. P. Quantitative proteomic analysis of amniocytes reveals potentially dysregulated molecular networks in Down syndrome. Clin Proteomics 2013, 10, 2, 10.1186/1559-0275-10-2doi

(74) Konvalinka, A.; Batruch, I.; Tokar, T.; Dimitromanolakis, A.; Reid, S.; Song, X.; Pei, Y.; Drabovich, A. P.; Diamandis, E. P.; Jurisica, I.; Scholey, J. W. Quantification of angiotensin II-regulated proteins in urine of patients with polycystic and other chronic kidney diseases by selected reaction monitoring. Clin Proteomics 2016, 13, 16, 10.1186/s12014-016-9117-x

(75) Drabovich, A. P.; Martinez-Morillo, E.; Diamandis, E. P. Toward an integrated pipeline for protein biomarker development. Biochim Biophys Acta 2015, 1854, 677–686, 10.1016/j.bbapap.2014.09.006

(76) Cho, C. K.; Drabovich, A. P.; Batruch, I.; Diamandis, E. P. Verification of a biomarker discovery approach for detection of Down syndrome in amniotic fluid via multiplex selected reaction monitoring (SRM) assay. J Proteomics 2011, 74, 2052–2059, 10.1016/j.jprot.2011.05.025

(77) Drabovich, A. P.; Jarvi, K.; Diamandis, E. P. Verification of male infertility biomarkers in seminal plasma by multiplex selected reaction monitoring assay. Mol Cell Proteomics 2011, 10, M110 004127, 10.1074/mcp.M110.004127

(78) Martinez-Morillo, E.; Cho, C. K.; Drabovich, A. P.; Shaw, J. L.; Soosaipillai, A.; Diamandis, E. P. Development of a multiplex selected reaction monitoring assay for quantification of biochemical markers of down syndrome in amniotic fluid samples. J Proteome Res 2012, 11, 3880–3887, 10.1021/pr300355a

(79) Prakash, A.; Rezai, T.; Krastins, B.; Sarracino, D.; Athanas, M.; Russo, P.; Ross, M. M.; Zhang, H.; Tian, Y.; Kulasingam, V.; Drabovich, A. P.; Smith, C. et al. Platform for establishing interlaboratory reproducibility of selected reaction monitoring-based mass spectrometry peptide assays. J Proteome Res 2010, 9, 6678–6688, 10.1021/pr100821m

(80) Prakash, A.; Rezai, T.; Krastins, B.; Sarracino, D.; Athanas, M.; Russo, P.; Zhang, H.; Tian, Y.; Li, Y.; Kulasingam, V.; Drabovich, A.; Smith, C. R. et al. Interlaboratory reproducibility of selective reaction monitoring assays using multiple upfront analyte enrichment strategies. J Proteome Res 2012, 11, 3986–3995, 10.1021/pr300014s

(81) Yang, W. L.; Lu, Z.; Guo, J.; Fellman, B. M.; Ning, J.; Lu, K. H.; Menon, U.; Kobayashi, M.; Hanash, S. M.; Celestino, J.; Skates, S. J.; Bast, R. C., Jr. Human epididymis protein 4 antigen-autoantibody complexes complement cancer antigen 125 for detecting early-stage ovarian cancer. Cancer 2020, 126, 725–736, 10.1002/cncr.32582

(82) Wang, X.; Yu, J.; Sreekumar, A.; Varambally, S.; Shen, R.; Giacherio, D.; Mehra, R.; Montie, J. E.; Pienta, K. J.; Sanda, M. G.; Kantoff, P. W.; Rubin, M. A. et al. Autoantibody signatures in prostate cancer. N Engl J Med 2005, 353, 1224–1235, 10.1056/NEJMoa051931

(83) Chen, W. S.; Haynes, W. A.; Waitz, R.; Kamath, K.; Vega-Crespo, A.; Shrestha, R.; Zhang, M.; Foye, A.; Baselga Carretero, I.; Perez Garcilazo, I.; Zhang, M.; Zhao, S. G. et al. Autoantibody Landscape in Patients with Advanced Prostate Cancer. Clin Cancer Res 2020, 26, 6204–6214, 10.1158/1078-0432.CCR-20-1966

(84) Meyer, S.; Woodward, M.; Hertel, C.; Vlaicu, P.; Haque, Y.; Karner, J.; Macagno, A.; Onuoha, S. C.; Fishman, D.; Peterson, H.; Metskula, K.; Uibo, R. et al. AIRE-Deficient Patients Harbor Unique High-Affinity Disease-Ameliorating Autoantibodies. Cell 2016, 166, 582–595, 10.1016/j.cell.2016.06.024

(85) Mintz, P. J.; Rietz, A. C.; Cardo-Vila, M.; Ozawa, M. G.; Dondossola, E.; Do, K. A.; Kim, J.; Troncoso, P.; Logothetis, C. J.; Sidman, R. L.; Pasqualini, R.; Arap, W. Discovery and horizontal follow-up of an autoantibody signature in human prostate cancer. Proc Natl Acad Sci U S A 2015, 112, 2515–2520, 10.1073/pnas.1500097112

(86) Reis, B. S.; Jungbluth, A. A.; Frosina, D.; Holz, M.; Ritter, E.; Nakayama, E.; Ishida, T.; Obata, Y.; Carver, B.; Scher, H.; Scardino, P. T.; Slovin, S. et al. Prostate cancer progression correlates with increased humoral immune response to a human endogenous retrovirus GAG protein. Clin Cancer Res 2013, 19, 6112–6125, 10.1158/1078-0432.CCR-12-3580

(87) Sreekumar, A.; Laxman, B.; Rhodes, D. R.; Bhagavathula, S.; Harwood, J.; Giacherio, D.; Ghosh, D.; Sanda, M. G.; Rubin, M. A.; Chinnaiyan, A. M. Humoral immune response to alpha-methylacyl-CoA racemase and prostate cancer. J Natl Cancer Inst 2004, 96, 834–843, 10.1093/jnci/djh145

(88) Arredouani, M. S.; Lu, B.; Bhasin, M.; Eljanne, M.; Yue, W.; Mosquera, J. M.; Bubley, G. J.; Li, V.; Rubin, M. A.; Libermann, T. A.; Sanda, M. G. Identification of the transcription factor single-minded homologue 2 as a potential biomarker and immunotherapy target in prostate cancer. Clin Cancer Res 2009, 15, 5794–5802, 10.1158/1078-0432.CCR-09-0911

(89) Lou, N.; Zheng, C.; Wang, Y.; Liang, C.; Tan, Q.; Luo, R.; Zhang, L.; Xie, T.; Shi, Y.; Han, X. Identification of novel serological autoantibodies in Chinese prostate cancer patients using high-throughput protein arrays. Cancer Immunol Immunother 2023, 72, 235–247, 10.1007/s00262-022-03242-0

(90) Fossa, A.; Berner, A.; Fossa, S. D.; Hernes, E.; Gaudernack, G.; Smeland, E. B. NY-ESO-1 protein expression and humoral immune responses in prostate cancer. Prostate 2004, 59, 440–447, 10.1002/pros.20025

(91) Westdorp, H.; Creemers, J. H. A.; van Oort, I. M.; Schreibelt, G.; Gorris, M. A. J.; Mehra, N.; Simons, M.; de Goede, A. L.; van Rossum, M. M.; Croockewit, A. J.; Figdor, C. G.; Witjes, J. A. et al. Blood-derived dendritic cell vaccinations induce immune responses that correlate with clinical outcome in patients with chemo-naive castration-resistant prostate cancer. J Immunother Cancer 2019, 7, 302, 10.1186/s40425-019-0787-6

(92) Chiriva-Internati, M.; Yu, Y.; Mirandola, L.; D’Cunha, N.; Hardwicke, F.; Cannon, M. J.; Cobos, E.; Kast, W. M. Identification of AKAP-4 as a new cancer/testis antigen for detection and immunotherapy of prostate cancer. Prostate 2012, 72, 12–23, 10.1002/pros.21400

(93) Rigamonti, N.; Bellone, M. Prostate cancer, tumor immunity and a renewed sense of optimism in immunotherapy. Cancer Immunol Immunother 2012, 61, 453–468, 10.1007/s00262-012-1216-6

(94) Tsai, C. T.; Robinson, P. V.; Spencer, C. A.; Bertozzi, C. R. Ultrasensitive Antibody Detection by Agglutination-PCR (ADAP). ACS Cent Sci 2016, 2, 139–147, 10.1021/acscentsci.5b00340

(95) Xie, C.; Kim, H. J.; Haw, J. G.; Kalbasi, A.; Gardner, B. K.; Li, G.; Rao, J.; Chia, D.; Liong, M.; Punzalan, R. R.; Marks, L. S.; Pantuck, A. J. et al. A novel multiplex assay combining autoantibodies plus PSA has potential implications for classification of prostate cancer from non-malignant cases. J Transl Med 2011, 9, 43, 10.1186/1479-5876-9-43

(96) Leidinger, P.; Keller, A.; Milchram, L.; Harz, C.; Hart, M.; Werth, A.; Lenhof, H. P.; Weinhausel, A.; Keck, B.; Wullich, B.; Ludwig, N.; Meese, E. Combination of Autoantibody Signature with PSA Level Enables a Highly Accurate Blood-Based Differentiation of Prostate Cancer Patients from Patients with Benign Prostatic Hyperplasia. PLoS One 2015, 10, e0128235, 10.1371/journal.pone.0128235

(97) Cottrell, T. R.; Lotze, M. T.; Ali, A.; Bifulco, C. B.; Capitini, C. M.; Chow, L. Q. M.; Cillo, A. R.; Collyar, D.; Cope, L.; Deutsch, J. S.; Dubrovsky, G.; Gnjatic, S. et al. Society for Immunotherapy of Cancer (SITC) consensus statement on essential biomarkers for immunotherapy clinical protocols. J Immunother Cancer 2025, *13*, 10.1136/jitc-2024-010928

(98) Mandl, S. J.; Rountree, R. B.; Dela Cruz, T. B.; Foy, S. P.; Cote, J. J.; Gordon, E. J.; Trent, E.; Delcayre, A.; Franzusoff, A. Elucidating immunologic mechanisms of PROSTVAC cancer immunotherapy. J Immunother Cancer 2014, 2, 34, 10.1186/s40425-014-0034-0

(99) Slovin, S. F.; Dorff, T. B.; Falchook, G. S.; Wei, X. X.; Gao, X.; McKay, R. R.; Oh, D. Y.; Wibmer, A. G.; Spear, M. A.; McCaigue, J.; Shedlock, D. J.; Dhar, M. et al. Phase 1 study of P-PSMA-101 CAR-T cells in patients with metastatic castration-resistant prostate cancer (mCRPC). Journal of Clinical Oncology 2022, 40, 98–98, 10.1200/JCO.2022.40.6_suppl.098

(100) Narayan, V.; Barber-Rotenberg, J. S.; Jung, I. Y.; Lacey, S. F.; Rech, A. J.; Davis, M. M.; Hwang, W. T.; Lal, P.; Carpenter, E. L.; Maude, S. L.; Plesa, G.; Vapiwala, N. et al. PSMA-targeting TGFbeta-insensitive armored CAR T cells in metastatic castration-resistant prostate cancer: a phase 1 trial. Nat Med 2022, 28, 724–734, 10.1038/s41591-022-01726-1

(101) Wargowski, E.; Johnson, L. E.; Eickhoff, J. C.; Delmastro, L.; Staab, M. J.; Liu, G.; McNeel, D. G. Prime-boost vaccination targeting prostatic acid phosphatase (PAP) in patients with metastatic castration-resistant prostate cancer (mCRPC) using Sipuleucel-T and a DNA vaccine. J Immunother Cancer 2018, 6, 21, 10.1186/s40425-018-0333-y

(102) Antonarakis, E. S.; Small, E. J.; Petrylak, D. P.; Quinn, D. I.; Kibel, A. S.; Chang, N. N.; Dearstyne, E.; Harmon, M.; Campogan, D.; Haynes, H.; Vu, T.; Sheikh, N. A. et al. Antigen-Specific CD8 Lytic Phenotype Induced by Sipuleucel-T in Hormone-Sensitive or Castration-Resistant Prostate Cancer and Association with Overall Survival. Clin Cancer Res 2018, 24, 4662–4671, 10.1158/1078-0432.CCR-18-0638

(103) Bhatia, V.; Kamat, N. V.; Pariva, T. E.; Wu, L. T.; Tsao, A.; Sasaki, K.; Sun, H.; Javier, G.; Nutt, S.; Coleman, I.; Hitchcock, L.; Zhang, A. et al. Targeting advanced prostate cancer with STEAP1 chimeric antigen receptor T cell and tumor-localized IL-12 immunotherapy. Nat Commun 2023, 14, 2041, 10.1038/s41467-023-37874-2

(104) Bakhshi, P.; Nourizadeh, M.; Sharifi, L.; Farajollahi, M. M.; Mohsenzadegan, M. Development of dendritic cell loaded MAGE-A2 long peptide; a potential target for tumor-specific T cell-mediated prostate cancer immunotherapy. Cancer Cell Int 2023, 23, 270, 10.1186/s12935-023-03108-0

(105) Daban, A.; Gonnin, C.; Phan, L.; Saldmann, A.; Granier, C.; Lillo-Lelouet, A.; Le Beller, C.; Pouchot, J.; Weiss, L.; Tartour, E.; Fabre, E.; Medioni, J. et al. Preexisting autoantibodies as predictor of immune related adverse events (irAEs) for advanced solid tumors treated with immune checkpoint inhibitors (ICIs). Oncoimmunology 2023, 12, 2204754, 10.1080/2162402X.2023.2204754

(106) Sato, Y.; Fujiwara, S.; Hata, A.; Kida, Y.; Masuda, T.; Amimoto, H.; Matsumoto, H.; Miyoshi, K.; Otsuka, K.; Tomii, K. Clinical Impact of Pre-Existing Autoantibodies in Patients With SCLC Treated With Immune Checkpoint Inhibitor: A Multicenter Prospective Observational Study. JTO Clin Res Rep 2023, 4, 100608, 10.1016/j.jtocrr.2023.100608

(107) Borgers, J. S. W.; van Wesemael, T. J.; Gelderman, K. A.; Rispens, T.; Verdegaal, E. M. E.; Moes, D.; Korse, C. M.; Kapiteijn, E.; Welters, M. J. P.; van der Burg, S. H.; van Houdt, W. J.; van Thienen, J. V. et al. Autoantibody-positivity before and seroconversion during treatment with anti-PD-1 is associated with immune-related adverse events in patients with melanoma. J Immunother Cancer 2024, *12*, 10.1136/jitc-2024-009215

(108) Pandit-Taskar, N.; O’Donoghue, J. A.; Durack, J. C.; Lyashchenko, S. K.; Cheal, S. M.; Beylergil, V.; Lefkowitz, R. A.; Carrasquillo, J. A.; Martinez, D. F.; Fung, A. M.; Solomon, S. B.; Gonen, M. et al. A Phase I/II Study for Analytic Validation of 89Zr-J591 ImmunoPET as a Molecular Imaging Agent for Metastatic Prostate Cancer. Clin Cancer Res 2015, 21, 5277–5285, 10.1158/1078-0432.CCR-15-0552

(109) Frigerio, B.; Morlino, S.; Luison, E.; Seregni, E.; Lorenzoni, A.; Satta, A.; Valdagni, R.; Bogni, A.; Chiesa, C.; Mira, M.; Canevari, S.; Alessi, A. et al. Anti-PSMA (124)I-scFvD2B as a new immuno-PET tool for prostate cancer: preclinical proof of principle. J Exp Clin Cancer Res 2019, 38, 326, 10.1186/s13046-019-1325-6

(110) Albano, D.; Piccardo, A.; Rizzo, A.; Cuzzocrea, M.; Bottoni, G.; Bellini, P.; Bertagna, F.; Treglia, G. Diagnostic performance of 2-[(18)F]FDG PET/CT in recurrent differentiated thyroid cancer and elevated antithyroglobulin antibodies: an updated systematic review and bivariate meta-analysis. Endocrine 2025, 87, 351–361, 10.1007/s12020-024-03989-9

