## Supporting Figures for "Identification and Quantification of Autoantibodies against Prostate-Specific Antigens by Immunoaffinity-Mass Spectrometry"

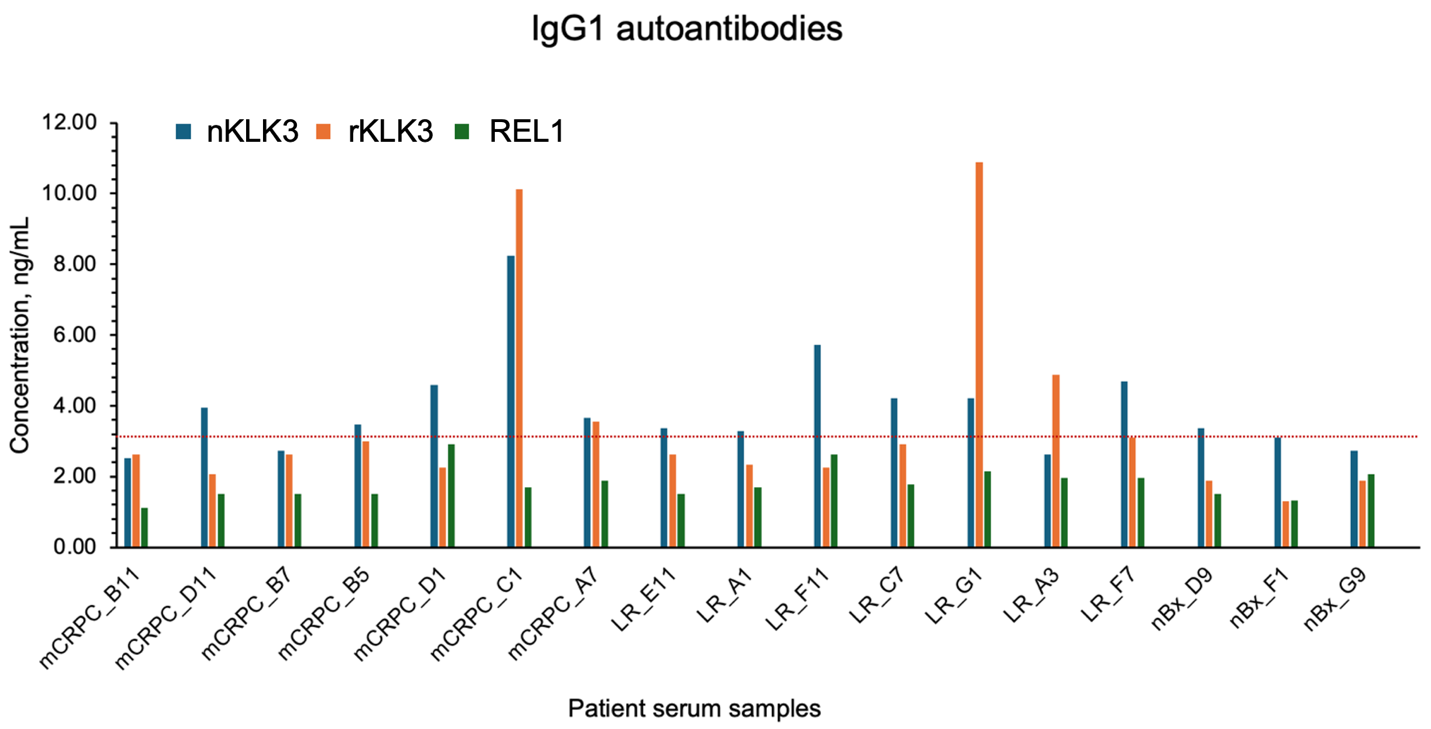

**Supplementary Figure S1****. Comparison of anti-PSA IgG1 enriched with recombinant PSA (rKLK3) and native PSA (nKLK3).** The concentration of anti-PSA IgG1 autoantibodies were found to correlate for most of the samples when measured against either rKLK3 or nKLK3. In this set of samples 3 nBx samples (F1, D9, and G9) were measured. The metastatic CRPC C1 (metC1) and A7 samples were confirmed to be positive for anti-PSA IgG1 autoantibodies against both antigens. The low-risk G1, and F7 samples were also detected positive (>LOD). However, the slight difference in concentration observed between the rKLK3 and nKLK3 might be associated to the difference in glycosylation.

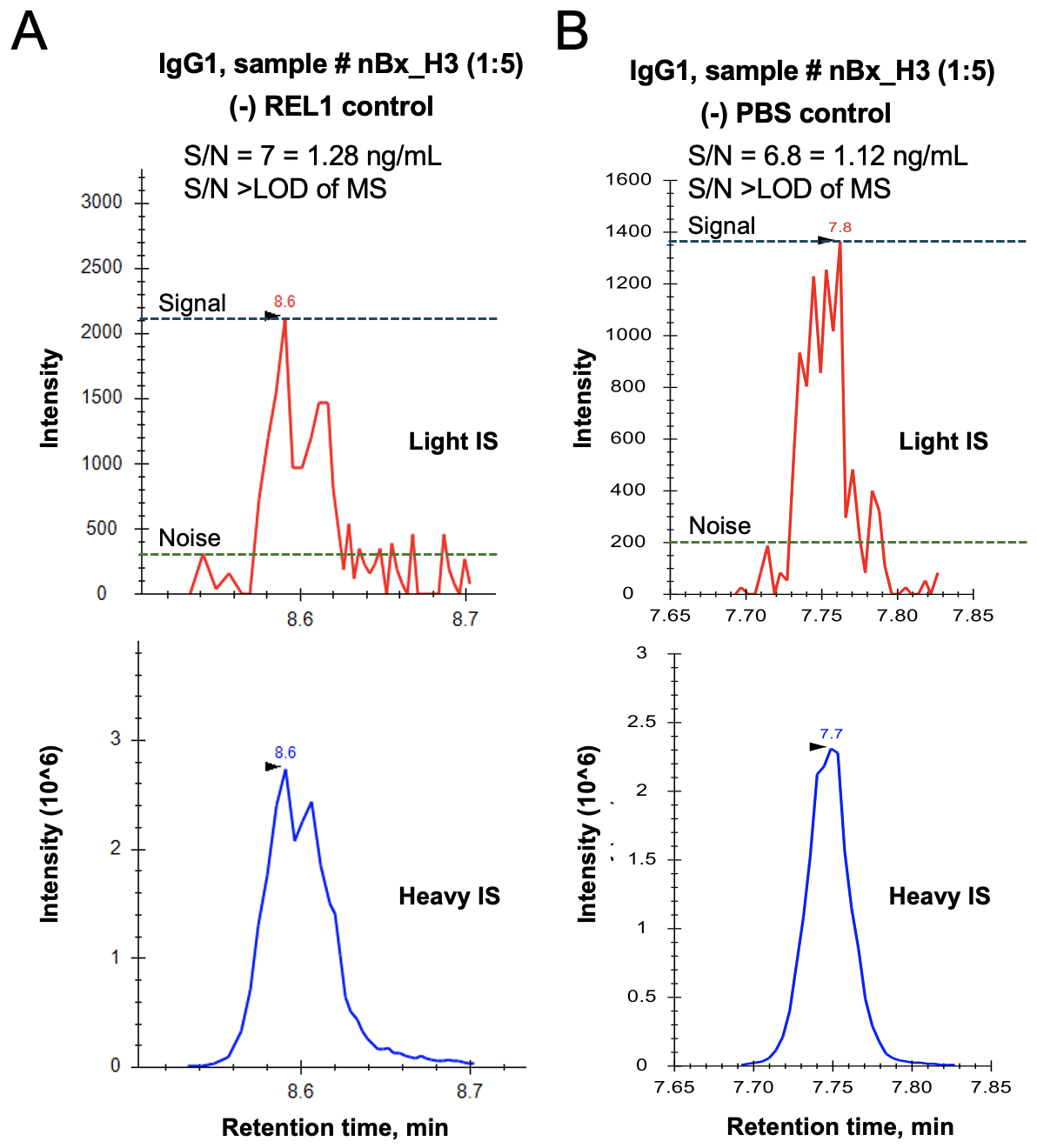

**Supplementary Figure S2. Determination of limit of detection (LOD) of IA-SRM assay.** Since each patient has a unique repertoire of polyclonal antibodies with potentially unique concentrations, epitopes, and affinities for each clone, a universal standard of antigen-specific polyclonal antibodies cannot be established and used to calculate assay LOD. Additional dilution of serum samples would unpredictable change the equilibrium established at 5-fold dilution of serum. (**A, B**) Thus, to estimate LOD, we calculated S/N in a sample with the lowest intensity of a light endogenous peptide (anti-REL1 and anti-PBS IgG1 in nBx_H3 sample). LOD was estimated at ~1 ng/mL.

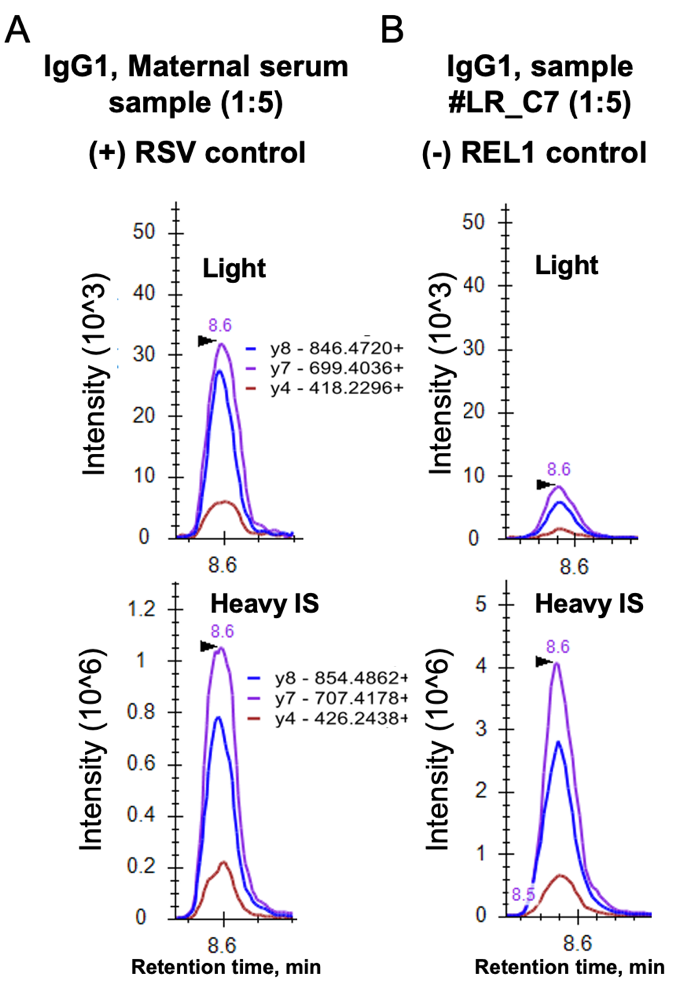

**Supplementary Figure S3. Representation of a positive and negative control used for the IA-SRM assays.** (**A**) Positive control showing the quantification of anti-RSV IgG1 in a maternal sera sample. (**B**) Negative control showing the quantification of anti-REL1 IgG1 in low risk C7 PCa serum sample. The positive control demonstrates a 3-fold higher intensity versus the negative control.

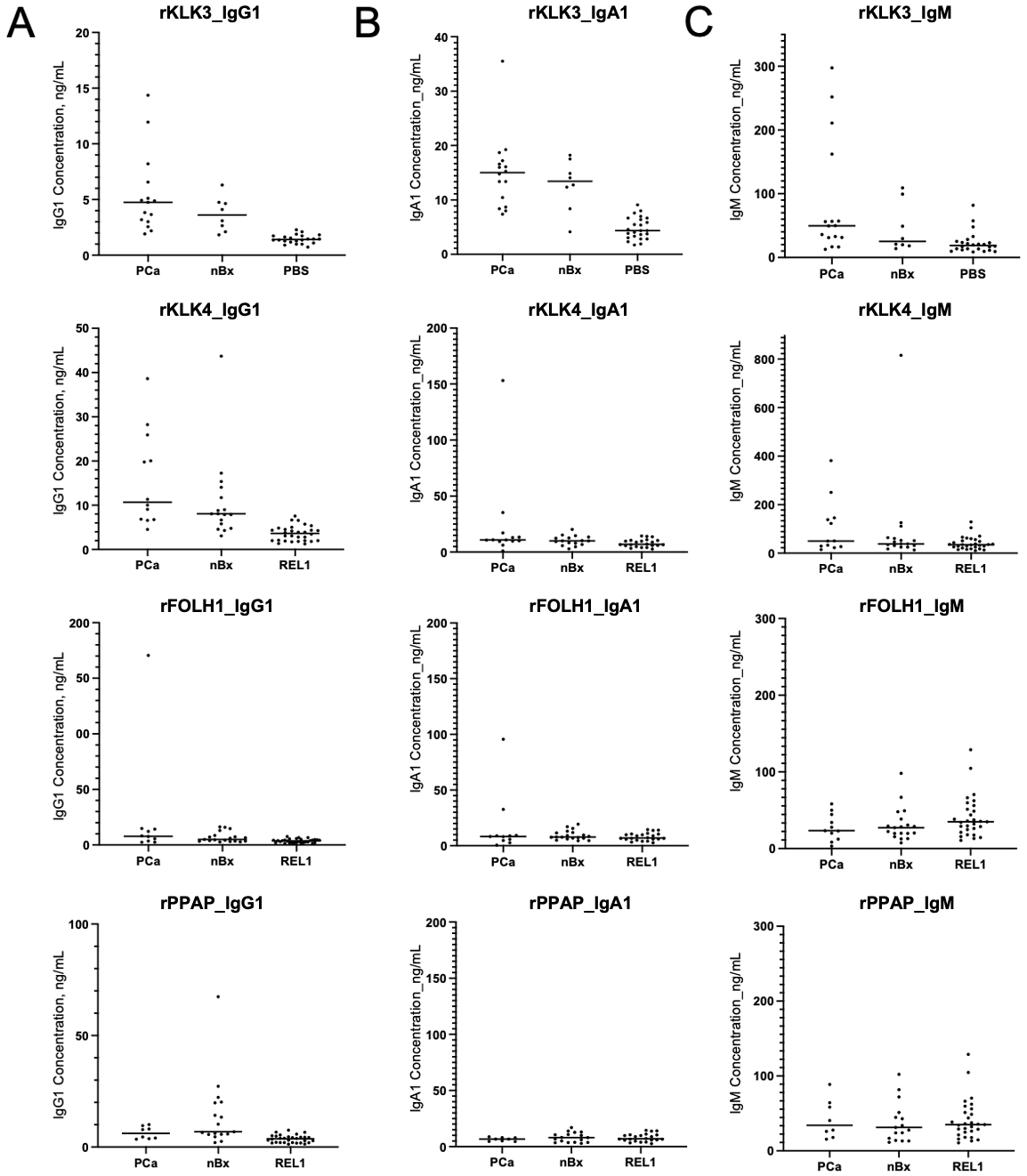

**Supplementary Figure S****4. Detection of anti-IgG1, -IgA1, and -IgM autoantibodies against PSA, KLK4, FOLH1, and PPAP antigens**. (**A, B**) Anti-IgG1 and anti-IgA1were detected in few negative biopsy and PCa samples against each of PSA, rKLK4, rPPAP, and rFOLH1 antigens. (**C**) Anti-IgM was found expressed at significantly high levels in PCa samples mainly against PSA and rKLK4. IA-MS assays enabled detection of much lower levels of rFOLH1-, or rPPAP-specific IgGM autoantibodies with higher background level.

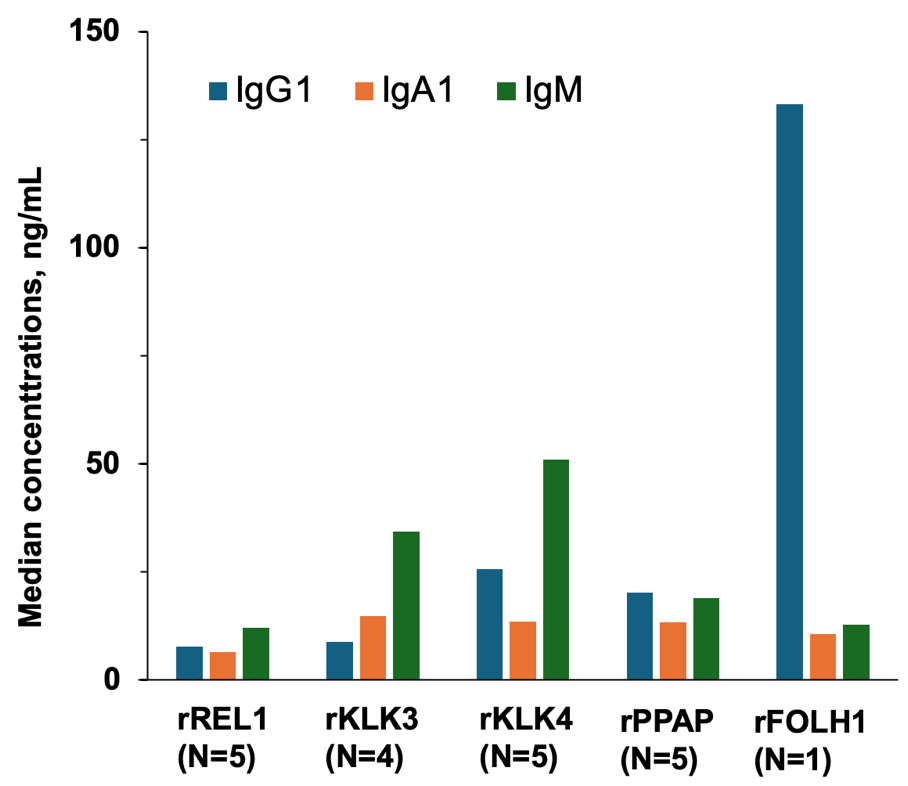

**Supplementary Figure S5. Medians concentrations of IgG1, IgA1 and IgM autoantibodies for each rKLK3, rKLK4, rPPAP, and rFOLH1 antigens.**

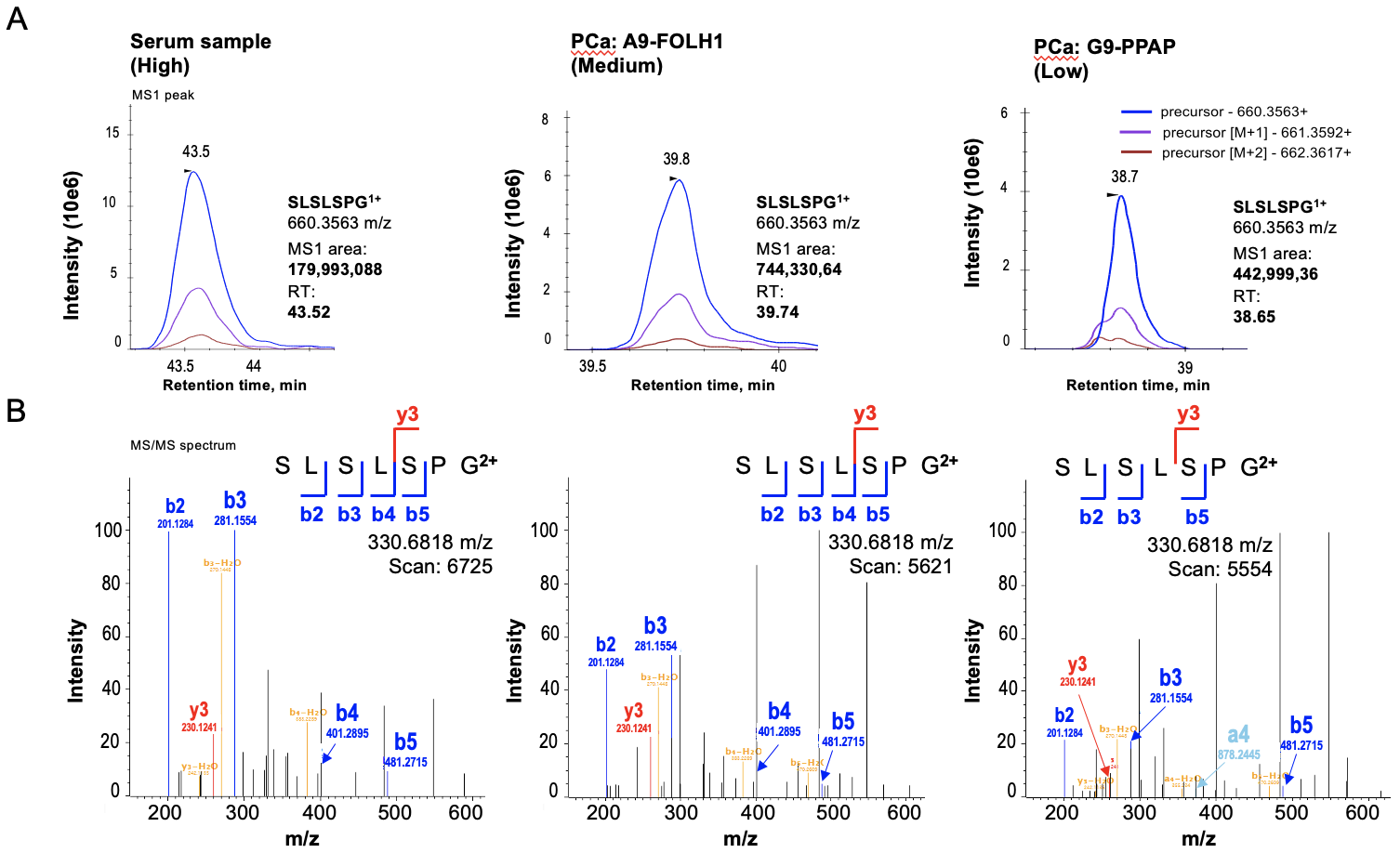

**Supplementary Figure S6. The secreted form of IgG1 autoantibodies detected using shotgun LC-MS/MS.** (**A**) MS1 of SLSLSPG peptide at +1 charge, the unique peptide of secreted IgG1, in samples expressing high, medium and low levels of IgG1. (**B**) MS/MS spectrum of SLSLSPG peptide at +2 charge. Fragments of SLSLSPG peptide were confirmed in the sample with a high level of IgG1 to be b2, b3, b4, b5 and y3 ions.
